# Dexamethasone modulate TWIST mediated EndMT changes in venous EC under acute shear stress. Implications for vein grafts disease

**DOI:** 10.1101/2023.05.26.23290570

**Authors:** Shameem Ladak, Liam W McQueen, Kristina Tomkova, Adewale Adebayo, Saadeh Suleiman, Sarah J George, Gavin J Murphy, Mustafa Zakkar

## Abstract

**Background:** The use of vein grafts in coronary artery surgery is complicated by a high late restenosis rate resulting from the development of intimal hyperplasia, and accelerated atherosclerosis. TGFβ has been implicated in the process of intimal hyperplasia but the role of TGFβ driven Endothelial to mesenchymal is not fully understood. Here, we have investigated the hypothesis that arterial shear stress (flow) can trigger Endothelial to mesenchymal changes in venous ECs mediated by TGFβ / SMAD pathway *in-vitro* and *ex-vivo* and that a brief pretreatment of vein with Dexamethasone can suppress such changes.

**Methods and Results:** Comparative reverse-transcriptase polymerase chain reaction, immunostaining and Western blotting revealed that arterial shear stress induced TGFβ / SMAD dependent in HUVEC which was regulated by TWIST 1&2 as the selective inhibition of TWIST 1 or 2 using specific siRNA suppressed EndMT in response to shear stress. We also noted that brief pretreatment of HUVECs with Dexamethasone can modulate EndMT changes in response to shear stress. Using spatial cell sequencing in human long saphenous vein segments exposed to acute arterial flow identified a cluster of cells that had both EC and SMC phenotypes where TWIST2 was significantly upregulated. We validated the untargeted spatial findings in segments of veins under acute arterial flow *ex-vivo* using comparative reverse-transcriptase polymerase chain reaction, immunostaining and RNAscope and observed that Dexamethasone can suppress EndMT changes in vein segments by suppressing TGFβ / SMAD/ TWIST1 &2.

**Conclusion:** Dexamethasone brief pretreatment can suppress EndMT changes triggered by acute exposure of long saphenous vein segments to arterial haemodynamics by modulating TGFβ / SMAD / TWIST1 &2 pathway.

## Introduction

The long saphenous vein (LSV) remains the most frequently used conduit in CABG and although arterial grafts may have better patency, however, recent studies questioned the superiority of arterial grafts compared to LSV. (1-3) The LSV also have the advantage of more reliable handling characteristics and availability. The use of LSV is however complicated by high failure rates due to the development of intimal hyperplasia (IH). It is accepted that IH is an inflammatory process that can be triggered as soon as the vein is harvested, yet the disease mechanisms regulating the pathophysiological changes involved still not fully understood. (3-5) Such processes include vascular inflammation leading to the activation of MAP kinases, endothelial expression of adhesion molecules and chemokines, and recruitment of inflammatory cells early after surgery. Endothelial-to-mesenchymal transition (EndMT) is the process by which endothelial cells lose their cell-specific markers and morphology and acquire a mesenchymal cell-like phenotype. This process is regulated by a complex orchestration of several signalling pathways and has been implicated in regulating cell phenotype adaptation during human vein graft remodeling after grafting veins into the arterial circulation with the TGFβ / SMAD signalling pathway playing a pivotal role in regulating this process in vein grafts. We previously showed that acute exposure of venous ECs (VECs) but not arterial ECs to arterial shear stress is associated with the activation or different proinflammatory mediators. (6, 7) Furthermore, it was arterial but not venous shear that activated such responses in VECs. (8) Glucocorticoids are known to regulate a variety of important cardiovascular, metabolic, and immunological functions. Glucocorticoids bind to the glucocorticoid receptor (GR) leading to its activation which in turn can influence gene expression by transactivation and transrepression. (9) Glucocorticoids can inhibit MAPK activities via the induction of MKP-1 which can inhibit transcriptional and posttranscriptional mechanisms controlled by the MAPK. (10, 11) Glucocorticoids as also known to regulate NF-kB in different complex methods including transcription and translocation regulated mechanisms. (12-14) Additionally, it is recognised that Glucocorticoids can significantly inhibit TGFβ production and modulate the activities of different members of the SMAD family and EndMT. (15-18) We and other have assessed the role of dexamethasone (Dex) on the modulation of vascular inflammation and IH in vein grafts and showed significant efficacy in suppressing inflammation and inhibiting IH. (7, 19)

In this study we aim to gain insight into the effects of arterial haemodynamics on EndMT activation in HUVECs *in-vitro* and LSV *ex-vivo* and we explore the role of brief pretreatment with Dex in regulating EndMT in LSV.

## Methods

### Reagents and Antibodies

Pharmacological inhibitors of TGFβ (Selleckchem S21, SB505124), Dexamethasone (Sigma-Aldrich, D2915) and antibodies (Table 1.1) were purchased commercially.

**Table 1.1:**
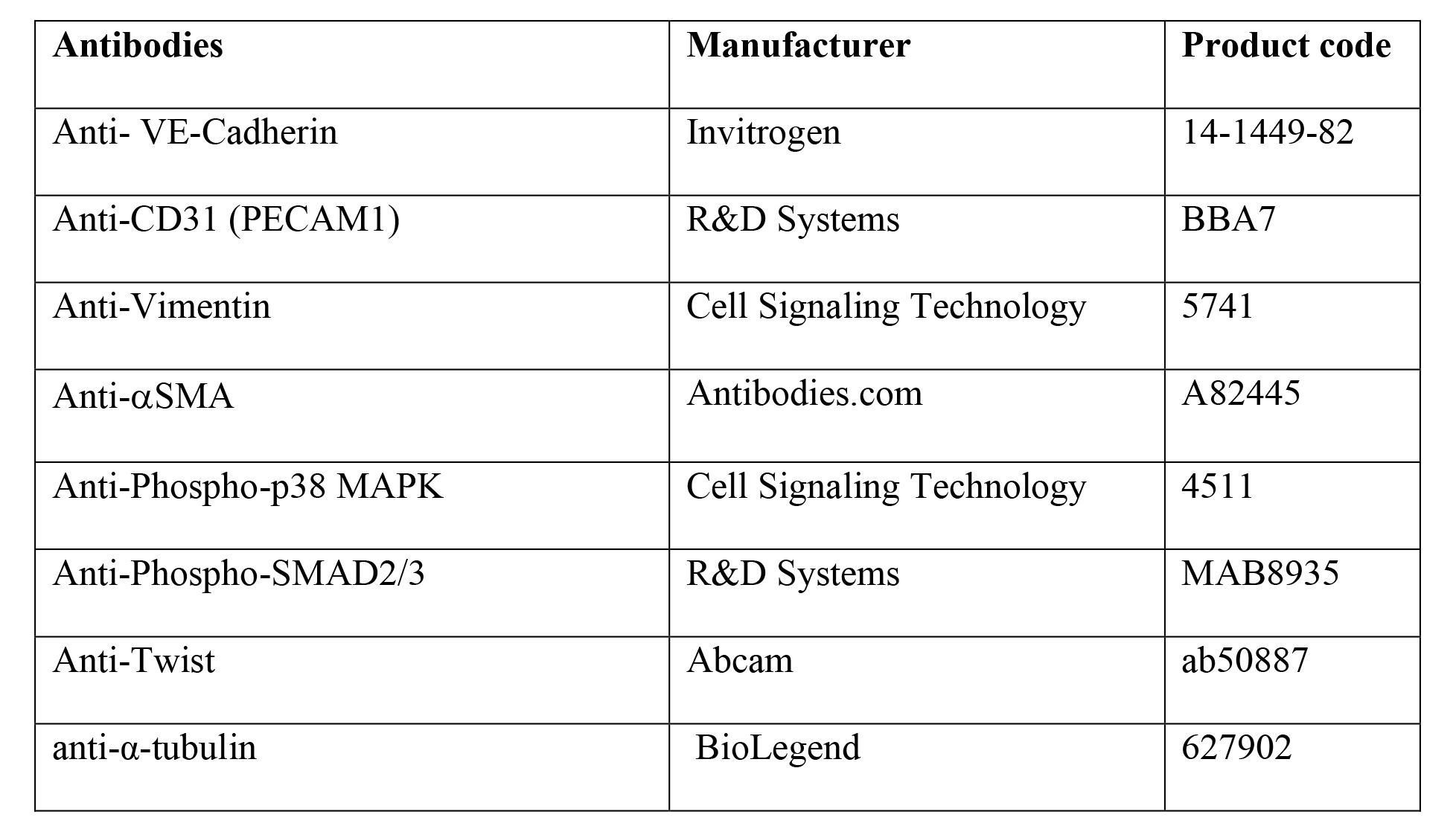
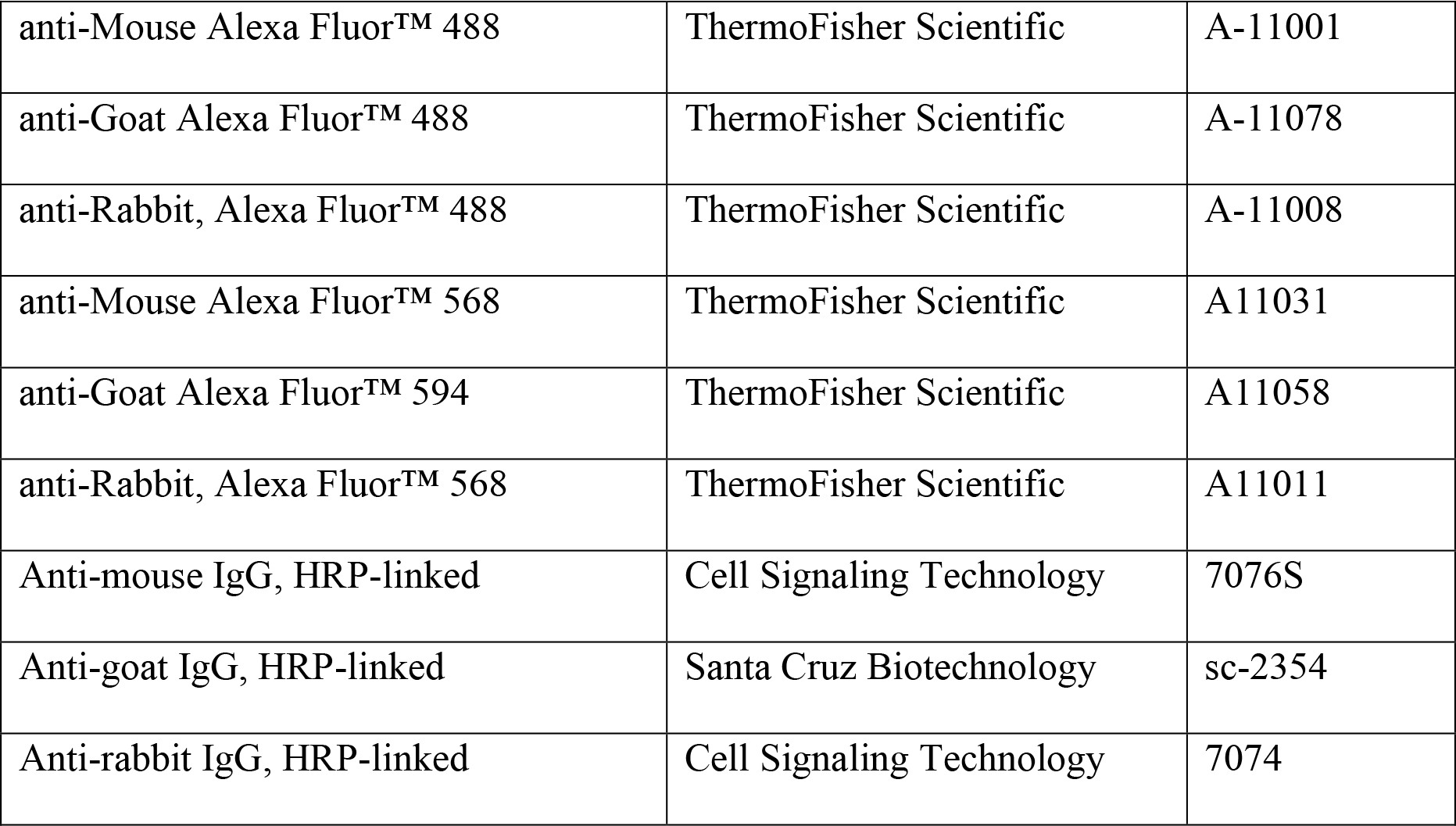
Antibodies

| Antibodies | Manufacturer | Product code |
| --- | --- | --- |
| Anti- VE-Cadherin | Invitrogen | 14-1449-82 |
| Anti-CD31 (PECAM1) | R&D Systems | BBA7 |
| Anti-Vimentin | Cell Signaling Technology | 5741 |
| Anti-αSMA | Antibodies.com | A82445 |
| Anti-Phospho-p38 MAPK | Cell Signaling Technology | 4511 |
| Anti-Phospho-SMAD2/3 | R&D Systems | MAB8935 |
| Anti-Twist | Abcam | ab50887 |
| anti-α-tubulin | BioLegend | 627902 |
| anti-Mouse Alexa Fluor™ 488 | ThermoFisher Scientific | A-11001 |
| anti-Goat Alexa Fluor™ 488 | ThermoFisher Scientific | A-11078 |
| anti-Rabbit, Alexa Fluor™ 488 | ThermoFisher Scientific | A-11008 |
| anti-Mouse Alexa Fluor™ 568 | ThermoFisher Scientific | A11031 |
| anti-Goat Alexa Fluor™ 594 | ThermoFisher Scientific | A11058 |
| anti-Rabbit, Alexa Fluor™ 568 | ThermoFisher Scientific | A11011 |
| Anti-mouse IgG, HRP-linked | Cell Signaling Technology | 7076S |
| Anti-goat IgG, HRP-linked | Santa Cruz Biotechnology | sc-2354 |
| Anti-rabbit IgG, HRP-linked | Cell Signaling Technology | 7074 |

### Comparative Reverse-Transcriptase Polymerase Chain Reaction

Total RNA was extracted using RNeasy mini kit (Qiagen, 74104). For mRNA studies cDNA was synthesized from total RNA using the Tetro cDNA synthesis kit (Bioline, BIO-65043). Gene expression was determined by quantitative real-time polymerase chain reaction (qRT-PCR) using SensiFast Probe Hi-ROX kit (Bioline, BIO-82005) and gene specific primers (Table 1.2) on Rotor gene Q (Qiagen) using manufacturer’s protocol. Relative levels were calculated using the 2^−(ΔΔCt)^ method and mRNA expression was normalized to housekeeping gene PPIA.

**Table 1.2:**
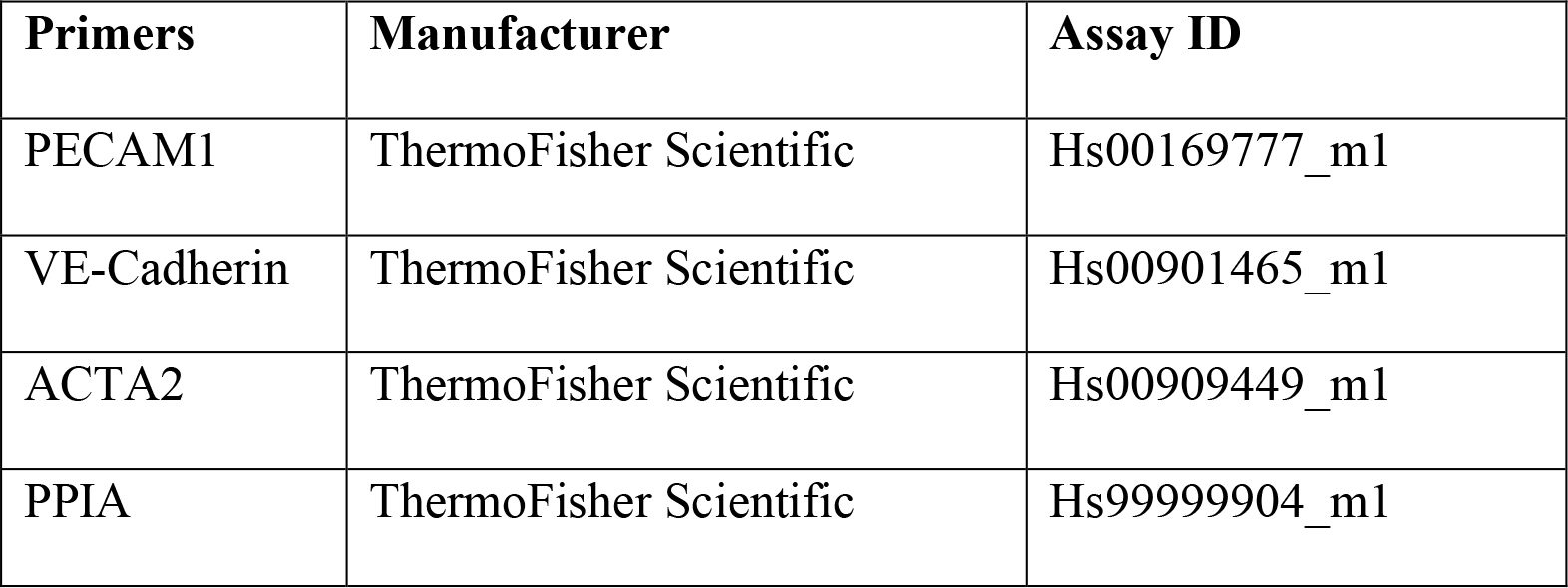
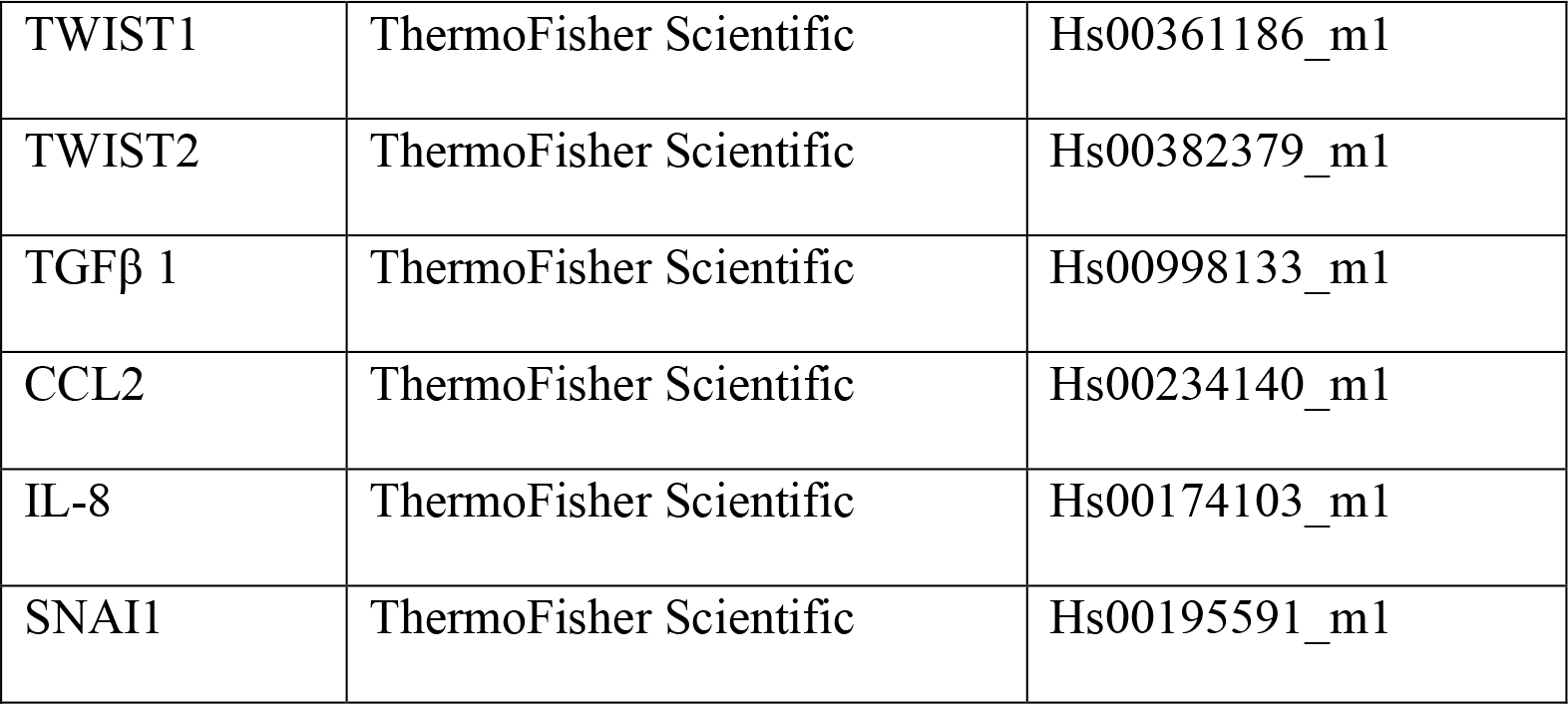
Primer list

| Primers | Manufacturer | Assay ID |
| --- | --- | --- |
| PECAM1 | ThermoFisher Scientific | Hs00169777_m1 |
| VE-Cadherin | ThermoFisher Scientific | Hs00901465_m1 |
| ACTA2 | ThermoFisher Scientific | Hs00909449_m1 |
| PPIA | ThermoFisher Scientific | Hs99999904_m1 |
| TWIST1 | ThermoFisher Scientific | Hs00361186_m1 |
| TWIST2 | ThermoFisher Scientific | Hs00382379_m1 |
| TGFβ 1 | ThermoFisher Scientific | Hs00998133_m1 |
| CCL2 | ThermoFisher Scientific | Hs00234140_m1 |
| IL-8 | ThermoFisher Scientific | Hs00174103_m1 |
| SNAI1 | ThermoFisher Scientific | Hs00195591_m1 |

### Silencing of TWIST-1 & TWIST-2

HUVECs were transiently transfected at 80% confluency using Lipofectamine RNAiMAX (Invitrogen, 13778075) transfection reagent and 30 μmol/L final concentration of TWIST1 siRNA (ThermoFisher Scientific, s14523) or TWIST2 siRNA (ThermoFisher Scientific, s42174) or Silencer™ Select Negative Control No. 1 siRNA (ThermoFisher Scientific, 4390843) in Opti-MEM reduced serum media (ThermoFisher Scientific, 31985070) according to manufacturer’s protocol for 24 hrs. All *in-vitro* experiments were conducted using HUVECs grown until passage 7.

### EC culture and shear stress

Pooled HUVECs were obtained from PromoCell (C-12203) and were cultured in Endothelial Cell Growth medium 2 (PromoCell, C-22011) to full confluency on glass microscope slides precoated with 1% (v/v) gelatin (Sigma-Aldrich, G1393). HUVECs were then exposed to laminar, unidirectional shear stress (at 0.5 or 12 dyn/cm^2^ to simulate venous and arterial rates of shear stress, respectively) for varying times, using parallel plate flow chambers designed in-house (Supplementary figure 2), as described previously, or maintained in static conditions. Briefly, the glass slides were placed into the parallel plate chambers and sealed with a silicon sheet gasket. A reservoir containing 30mL RPMI 1640 culture medium (ThermoFisher Scientific 11875093), supplemented with 2% (v/v) FCS, 100µg/ml penicillin and 100U/ml streptomycin attached to a closed-circuit loop of silicon tubing (VWR, USA and Elkay, UK) was connected to the chambers. HUVECs were then cultured at 37°C and 5% CO_2_ and shear stress applied using a multi-channel peristaltic pump (Watson-Marlow, UK). Shear stress rates were calculated for a slit die assuming the viscosity of water at 37°C, using the following equation: τ=6µQ/wh^2^. Where τ represents Shear Stress; µ, viscosity; Q, flow rate; w, width and h, height.

### *Ex-vivo* perfusion of veins

Surplus segments of surgically prepared human long saphenous veins (LSV), resected during coronary artery bypass graft surgery from anonymised consenting patients, were placed immediately in RPMI 1640 culture medium with 10% (v/v) FCS, 100µg/ml penicillin and 100U/ml streptomycin. The study was approved under Leicester Biomedical Research Centre (BRICCS Ethics Ref: 09/H0406/114). Informed consent was obtained from all study participants prior to surgery and use of human tissue conformed to the principles outlined in the Declaration of Helsinki. Appropriate length sections were cut transversely for static/baseline control measurements and maintained in RPMI 1640 culture medium; 6cm sections were cut for use in *ex-vivo* perfusion. Vein sectprotein was extracted with RIPA ions were cannulated with Male Luer Fittings 1/16” (World Precision Instruments, USA) and secured with fine surgical tie. Secured veins were then attached to the flow system was placed in an incubator and maintained at 37°C. Veins were perfused under mean arterial pressure (65 mm Hg) with M199 medium (containing 20% FCS, 100 U/mL penicillin, and 0.1 mg/mL streptomycin), which was oxygenated (oxygen content, 20 mL/L) and prewarmed to 37°C. We targeted a wall shear stress of 0.5±0.1 dyn/cm^2^ for low flow experiments and 12±0.2dyn/cm^2^ for high shear and adjusted flow rates based on the diameter of the cannulae, shear stress through a non-deformable cylinder assuming laminar flow and the viscosity of water at 37°C, using the equation: τ=4µQ/πr^3^. Where τ represents Shear Stress; µ, viscosity; Q, flow rate; π, pi; r, radius.

### Immunoblotting

Total HUVEC protein was extracted with RIPA Lysis and Extraction Buffer lysis buffer (ThermoFisher Scientific, 89900), or, alternatively, cell compartment lysates were prepared with the Nuclear Extraction Kit (Active Motif, 40010). All SDS-Polyacrylamide Gel Electrophoresis (PAGE) were performed using the Bio-Rad Mini format 1-D electrophoresis system, 4-15% Mini-PROTEAN TGX stain free gels (Bio-Rad, 4561095) and 10x Tris/Glycine/SDS electrophoresis buffer (Bio-Rad, 1610772). Following transfer, 0.2µM Nitrocellulose membranes (Bio-Rad, 1620112), were blocked in 5% (w/v) semi-skimmed milk powder and primary antibodies (Table 1.1) were incubated overnight at 4°C. Primary antibodies were detected by Horseradish Peroxidase-conjugated secondary antibodies (Table 1.1) for 2 hrs at RT and SuperSignal™ West Pico PLUS Chemiluminescent Substrate kit (ThermoFisher Scientific). Membranes were imaged using the ImageQuantTL imaging system and detected protein bands were quantified by densitometry and normalised to the stain-free loading control (GAPDH).

### Immunocytochemistry

HUVECs cultured on glass slides (Fisher Scientific, 13192131) were exposed to shear stress or maintained under static conditions and fixed in slides using pre chilled 10% neutral buffered formalin (Sigma Aldrich, HT5014-1CS) for 15 mins at 4°C. Thereafter, slides were dehydrated in ethanol (50-100%) and allowed to dry. Cells were incubated in Immunofluorescence Blocking Buffer (Cell Signaling Technology, 12411) for 30 mins at room temperature followed by overnight primary antibody (Table 1.1) incubation in Immunofluorescence Antibody Dilution Buffer (Cell Signaling Technology, 12378) at 4°C and AlexaFluor fluorophore-conjugated secondary antibody (Table 1.1) for 1 hr at room temperature. Immunoglobulin matched controls at the same concentration as the primary antibodies were used as staining-specific negative controls. Nuclei were labelled with diamidino phenylindole (DAPI)-dilactate (Invitrogen) in PBS for 30 secs and coverslips were mounted with ProLong® Gold Antifade Reagent (ThermoFisher Scientific, P36930). Slides were visualized using Zeiss Axio Observer Z1 inverted microscope and quantification of fluorescence intensity was measured as integrated intensity in whole cells divided by total number of cells using CellProfiler 2.0

### LSV sections immunostaining

The expression levels of specific proteins were assessed in veins by immunostaining. For immunostaining on fresh frozen tissues, slides were incubated in pre chilled 10% neutral buffered formalin (Sigma Aldrich, HT5014-1CS) for 15 mins at 4°C. Thereafter, slides were dehydrated in ethanol (50-100%) and allowed to dry. For staining on formalin-fixed, paraffin-embedded human samples, slides were deparaffinized and underwent antigen retrieval. Sections were then incubated in Immunofluorescence Blocking Buffer (Cell Signalling Technology, 12411) for 30 mins at room temperature followed by overnight primary antibody (Table 1.1) incubation in Immunofluorescence Antibody Dilution Buffer (Cell Signalling Technology, 12378) at 4°C. Secondary antibody (HRP linked, Table 1.1) was applied for one hour at RT followed by nuclei staining with DAPI. Control slides were routinely stained in parallel by substituting IgG, or the specific IgG isotype. All images were acquired using either a Zeiss Axioscope with AxioVision V4.3 software, a Zeiss LSM 510 UV laser scanning confocal microscope (Carl Zeiss, GmBH). The expression of particular proteins was assessed by quantification of fluorescence intensity for multiple cells (at least 100 per site) as reported previously (7, 8).

### RNAscope in situ hybridisation

Frozen vein sections were processed for fluorescent in situ hybridization by RNAscope according to manufacturer’s guidelines. Genes examined in the vein sections were TWIST1 (ACDBio, 470291), TWIST2 (ACDBio, and SNAI1 (ACDBio, 560421-C2) and hybridization was performed using RNAscope® Multiplex Fluorescent Reagent Kit v2 (ACDBio 323100). These sections were co-stained with CD31 or VE-Cadherin or α-SMA antibodies using RNAscope® Multiplex Fluorescent v2 Assay combined with Immunofluorescence - Integrated Co-Detection Workflow (Advanced Cell Diagnostics) to study the co-expression of TWIST1 and SNAI1 gene in ECs and SMCs. Slides were cover slipped with ProLong™ Gold Antifade Mountant and stored at 4°C in the dark before imaging. TWIST1 and SNAI1 puncta were counted selectively in CD31+ cells using the cell-counter plugin in Fiji (20). After the RNAscope assay was performed, dots quantification was done based on the average number of dots per cell as previously described. (21)

### *Ex-vivo* monocyte adhesion

Following 30 minutes incubation with 10 µmol/L Dex and exposure to acute arterial flow for 4 h, LSV segments were dissected longitudinally, pinned with the luminal surface facing upwards and co-cultured, in static conditions, with 1 × 10^6^ Calcein AM-labelled (10 µmol/L) THP-1 cells for 15 min. Immediately after co-culture and washing, in situ adhered monocytes and LSV segments were imaged using fluorescence microscopy (Zeiss, Germany).

### Vein Culture well

LSV were cultured after exposure to 4 hours of arterial flow in a culture well as previously described. (22) Briefly, after flow exposure, the vein were cut longitudinally and pinned with the endothelial surface up in segments of 5 to 10 mm in length. Veins were cultured at 37°C in bicarbonate-buffered RPMI 1640 supplemented with 30% FBS unless otherwise stated under an atmosphere of 95% air and 5% CO_2_ For 10 days.

### Spatial cell sequencing

For sectioning in preparation of Visium, blocks were equilibrated to −18^0^ C, and 10 mm thick sections were mounted onto the active sequencing areas of the 10x Genomics Visium slides. Hemotoxylin and eosin staining was performed according to the 10x Genomics Visium protocol (www.10xgenomics.com/). Briefly, tissue sections are placed onto the Visium Gene Expression Slide, fixed with ice cold methanol, stained with haematoxylin and eosin (H&E) and imaged using Olympus FV1000 confocal microscope. Tissue permeabilization was done according to optimised protocol and reverse transcription (RT) was undertaken on a thermocycler to produce spatially barcoded, full-length cDNA from poly-adenylated mRNA on the slide. Second strand synthesis used the Second Strand Reagent, Primer, and Enzyme mix, followed by denaturation and transfer of the cDNA from each capture area to a corresponding tube for cDNA amplification and library construction (following cycle number determination and quality control). Cycle number determination was undertaken using the provided KAPA SYBR FAST qPCR Master Mix and cDNA Primers, whilst cDNA amplification was undertaken using the provided Amp Mix and cDNA Primers. This resulted in a solution of spatially barcoded, full-length amplified cDNA generated by PCR for subsequent library construction. Final quality control was determined using the Aligent Bioanalyser High Sensitivity Kit. Library construction was undertaken using a fixed proportion of the total cDNA (40uL), with enzymatic fragmentation and size selection utilised to optimise the amplicon size. End-repair, A-tailing, adaptor ligation and PCR are utilised to add P5, P7, i7 and i5 sample indexes, along with a TruSeq Read 2 primer sequence. Libraries were sequenced using PE150 on NovaSeq platform (Novogene). Spatial sequencing data was processed using the SpaceRanger v1.3.1 pipeline, aligned to the associated 10X Genomics Human Genome Reference GRCh38-2020-A. Brightfield images in high resolution tiff format were manually aligned using 10x Loupe Browser 6.4.1 software, to ensure accurate orientations of the expression data to the tissue image. Count data and image data for all samples were imported into to Seurat v4.0 (https://pubmed.ncbi.nlm.nih.gov/34062119/) for further analysis. Data was normalised using SCTransform (https://genomebiology.biomedcentral.com/articles/10.1186/s13059-019-1874-1), correcting for batch effects between different runs. All sample datasets were integrated for joint comparisons following the Seurat SCTransform integration workflow (https://www.cell.com/cell/fulltext/S0092-8674(19)30559-8), with 3000 integration features and all common genes between samples. Aggregated spot expression data from all samples was tested with muscatv1.6 (https://www.nature.com/articles/s41467-020-19894-4) in R v4.0.0 to better account for variation in spots in a multi-sample, multi-batch, multi-state experiment with group replicates. Unsupervised clustering was performed using the Seurat FindClusters function, and cluster identity was inferred using a combination unique gene marker identification and analysis of the top 30 most highly expressed genes in each grouping. Pathway analysis was undertaken for samples reaching significance (p < 0.05 adjusted) using the gprofiler package (23) and associated public datasets (KEGG, REAC, Wikipathways, Gene Ontology, biological processes, cellular components, and molecular functions. (24-27) Network analysis was performed using the Cytoscape application (version 3.9.1). (23) Connections were inferred between genes of interest by running a heat diffusion analysis with standard parameters. (28) Global network analysis was subsequently undertaken using default confidence and interaction parameters. Genes were visually quantified using a colour gradient scale, ranging from red to blue based on their LogFC value. Specific genes of interest were identified and subsetted from the global network to identify nearest neighbouring interactors from known pathways which demonstrated predicted a significant overlap and connections between these different genes and pathways involved, using diffusion analysis. (28)

### Statistics

For experiments where only two groups were analysed, data were subjected to a paired, two-tailed t-test. For experiments where more than two groups were analysed, a one-or two-way ANOVA was used depending on the number of independent variables, followed by post-hoc pairwise comparisons with Bonferroni correction for multiple comparisons. If datasets were large enough (for example for immocytochemical analyses, where 20 images per sample were analysed), normal distribution was assessed with the D’Agostino-Pearson test; all data assessed passed normality tests, as such, parametric analyses were appropriate. The cut-off value for statistical significance was 0.05. Data are presented as mean ± SEM. All statistical analysis was performed with GraphPad Prism 7.0.

## Results

### Acute arterial shear stress promotes EndMT in Venous ECs

We looked at the effect of acute arterial shear stress (ASS) on the activation of EndMT. We observed by quantitative RT-PCR that the application of ASS for 4 hours activated VECs, as shown by expression of the chemokines MCP-1 and CXCL8 as previously reported. (7, 8) However, we note a significant down-regulation of VEcad and up-regulation of VIM, TWIST 1 and TWIST 2 gene expression (Figure. 1A). CD31 and SMA showed similar trends but were not statistically significant (Supp Figure. 1A). These data suggest that ASS can results in the activation of EndMT which can be potentially regulated by TGFβ. To examine this hypothesis, we used a pharmacological inhibitor of TGFβ that is known to specifically inhibit TGFβR1 (ALK5) (SB505124) and observed that pretreatment of HUVECS with this inhibitor for 60 mins suppressed the induction of the phenotype changes noted above both at RNA (Figure. 1B) and protein levels at 4 hours which was associated with a more rapid (45 min) suppression on ALK5 activation and SMAD 2/3 phosphorylation (Figure. 1C). Thus, we conclude that venous ECs are sensitive to TGFβ -dependent induction of EndMT by ASS.

**Figure. 1.**
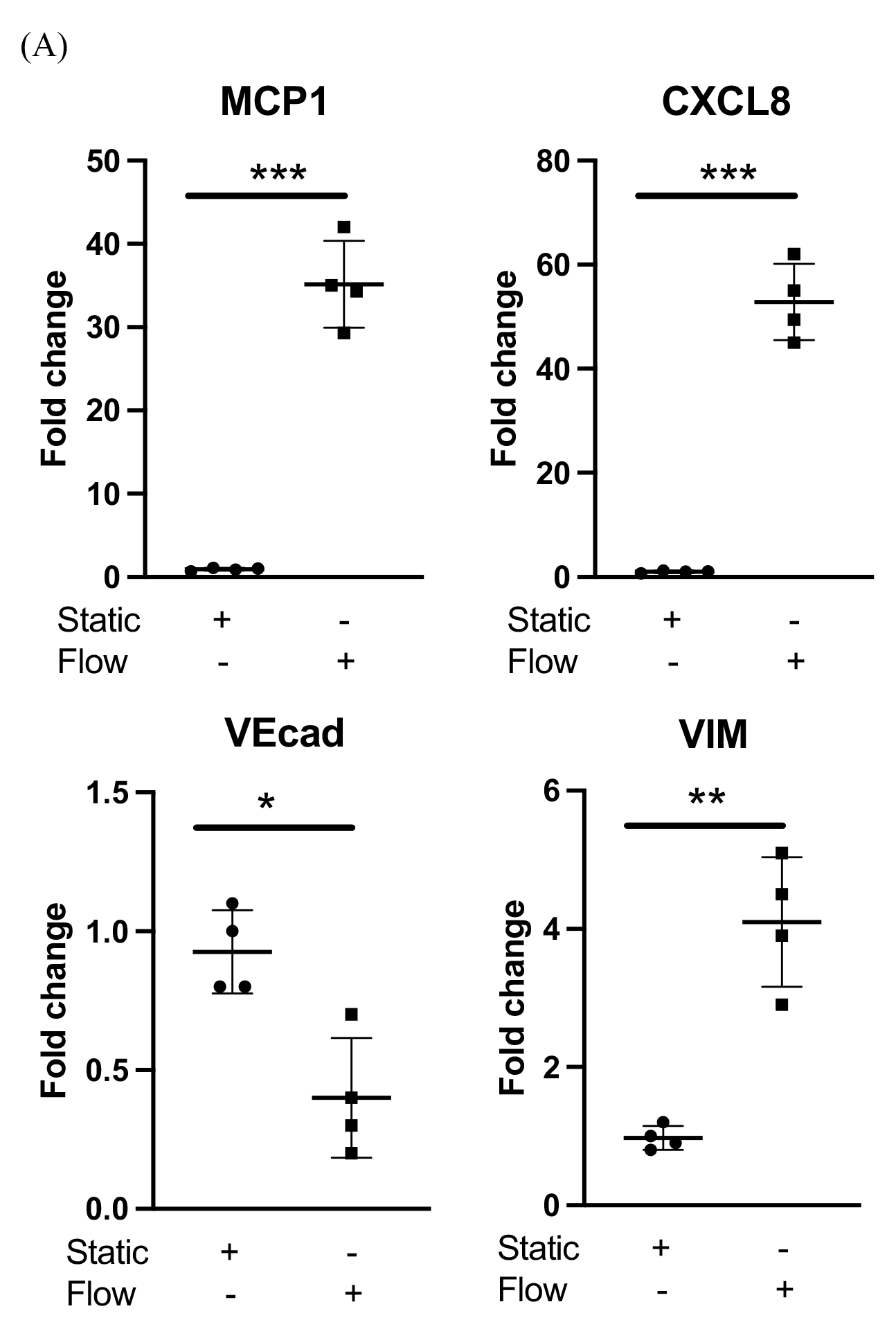
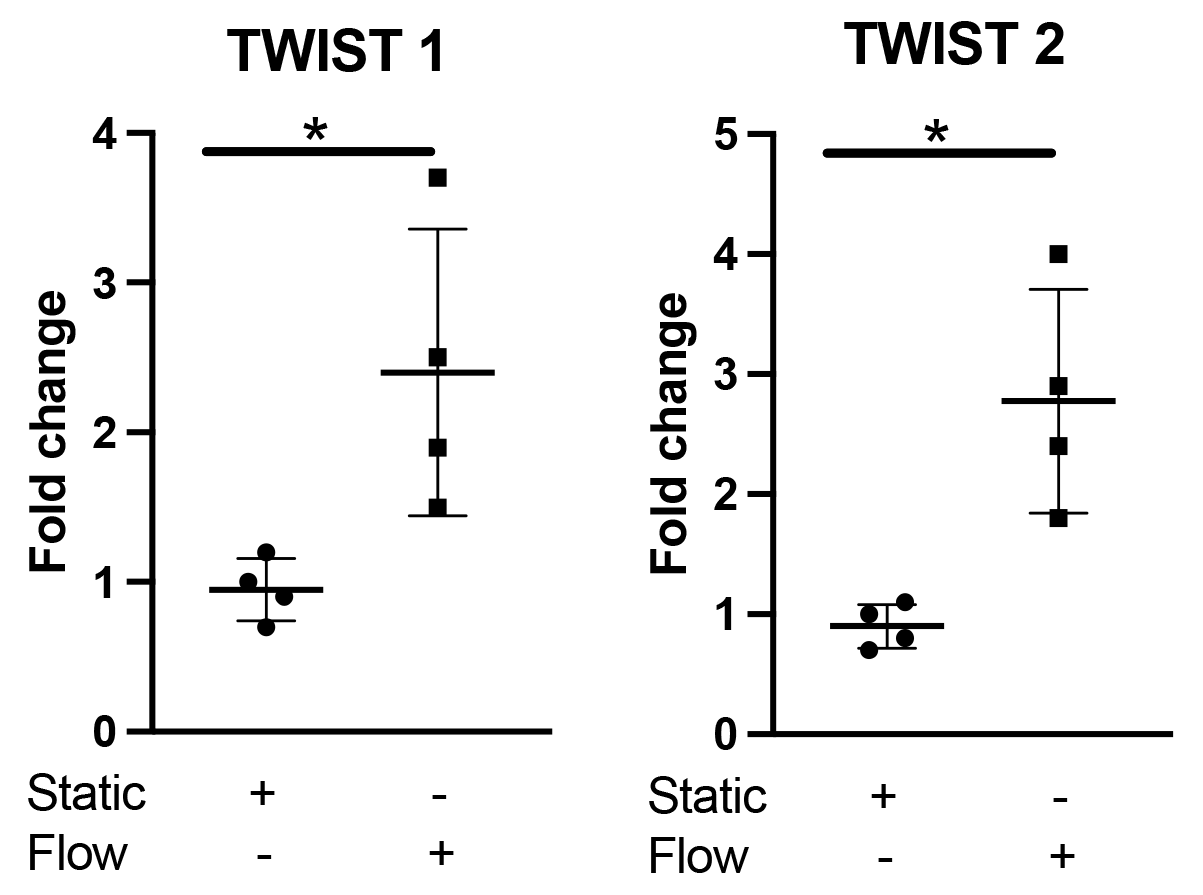
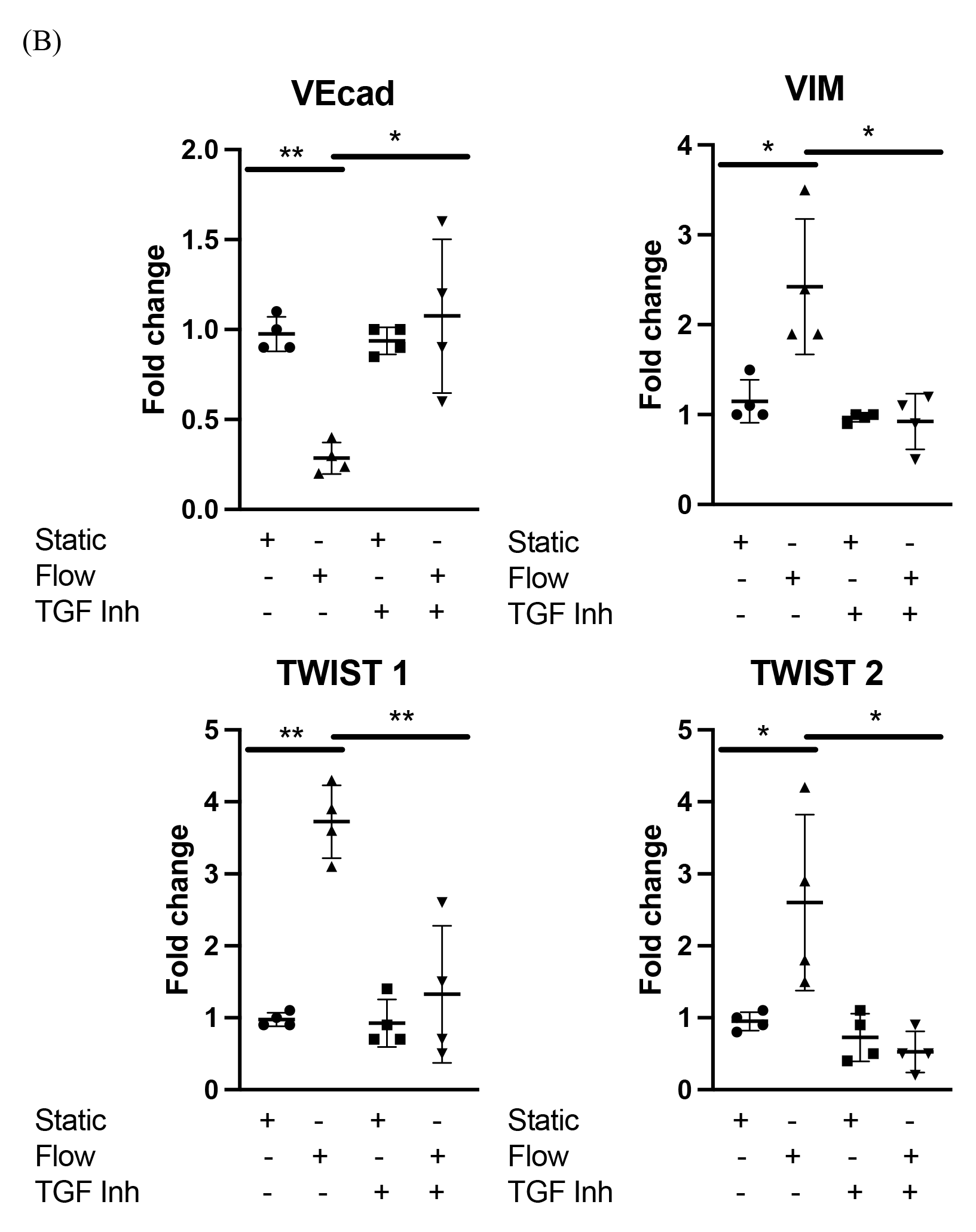
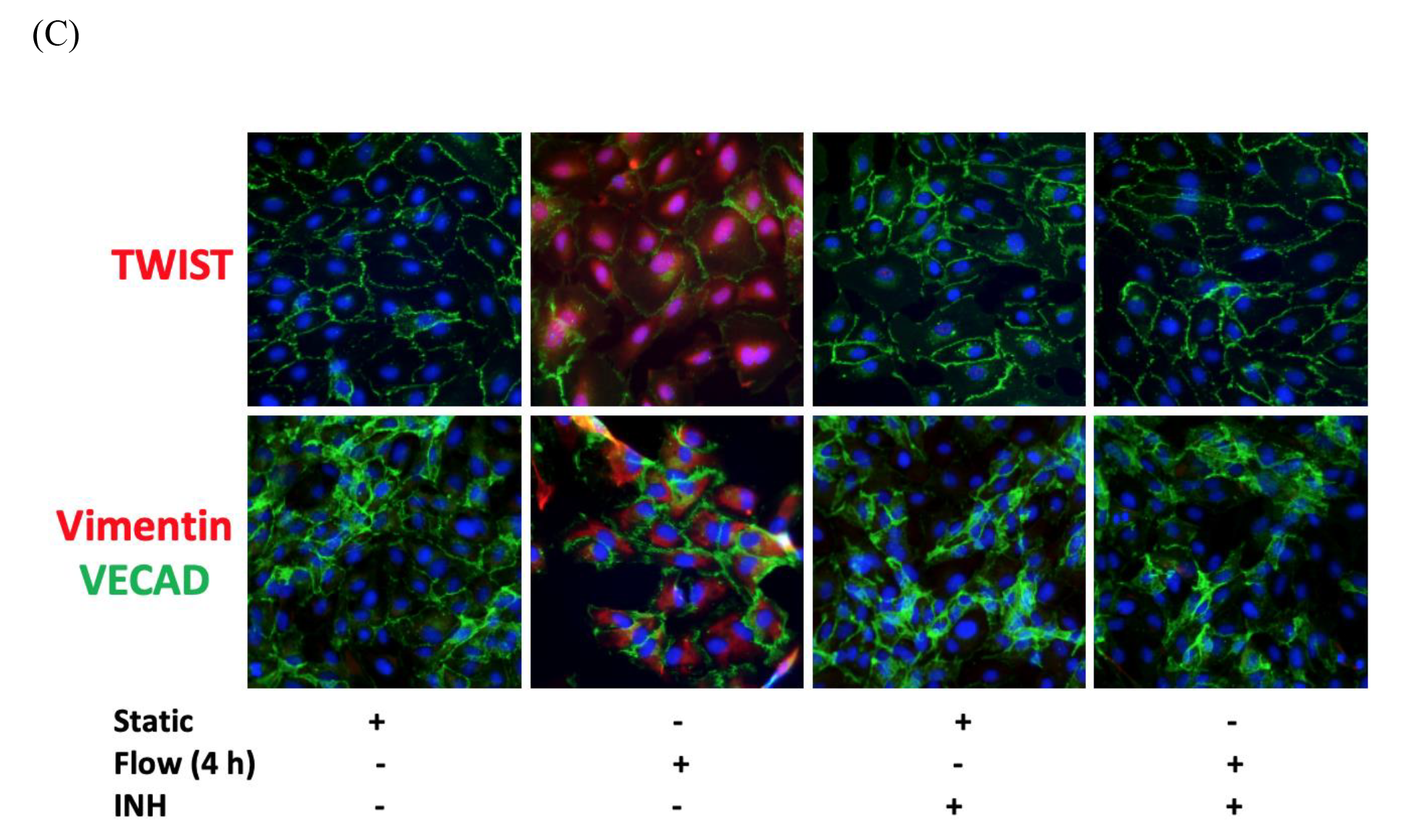
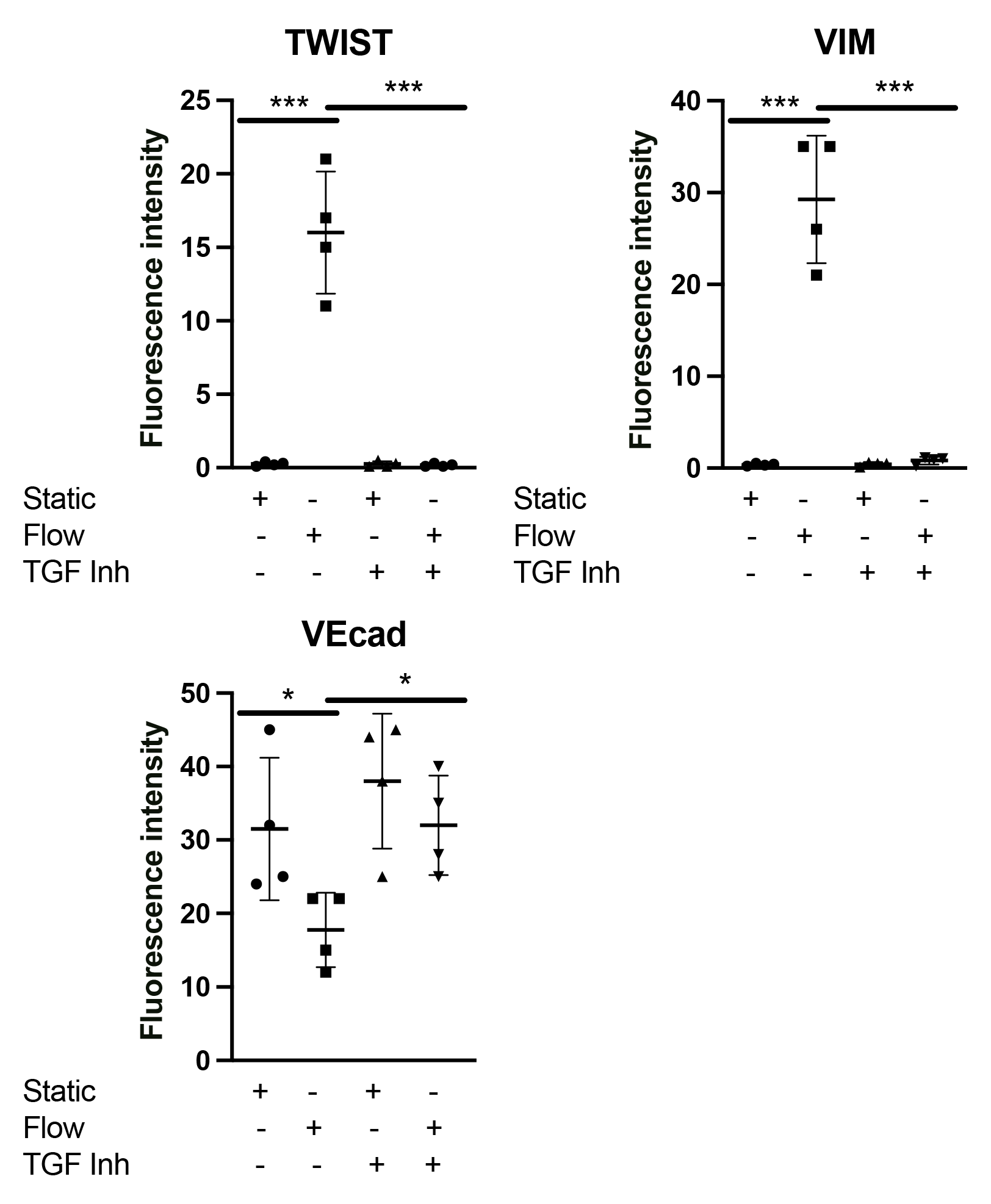
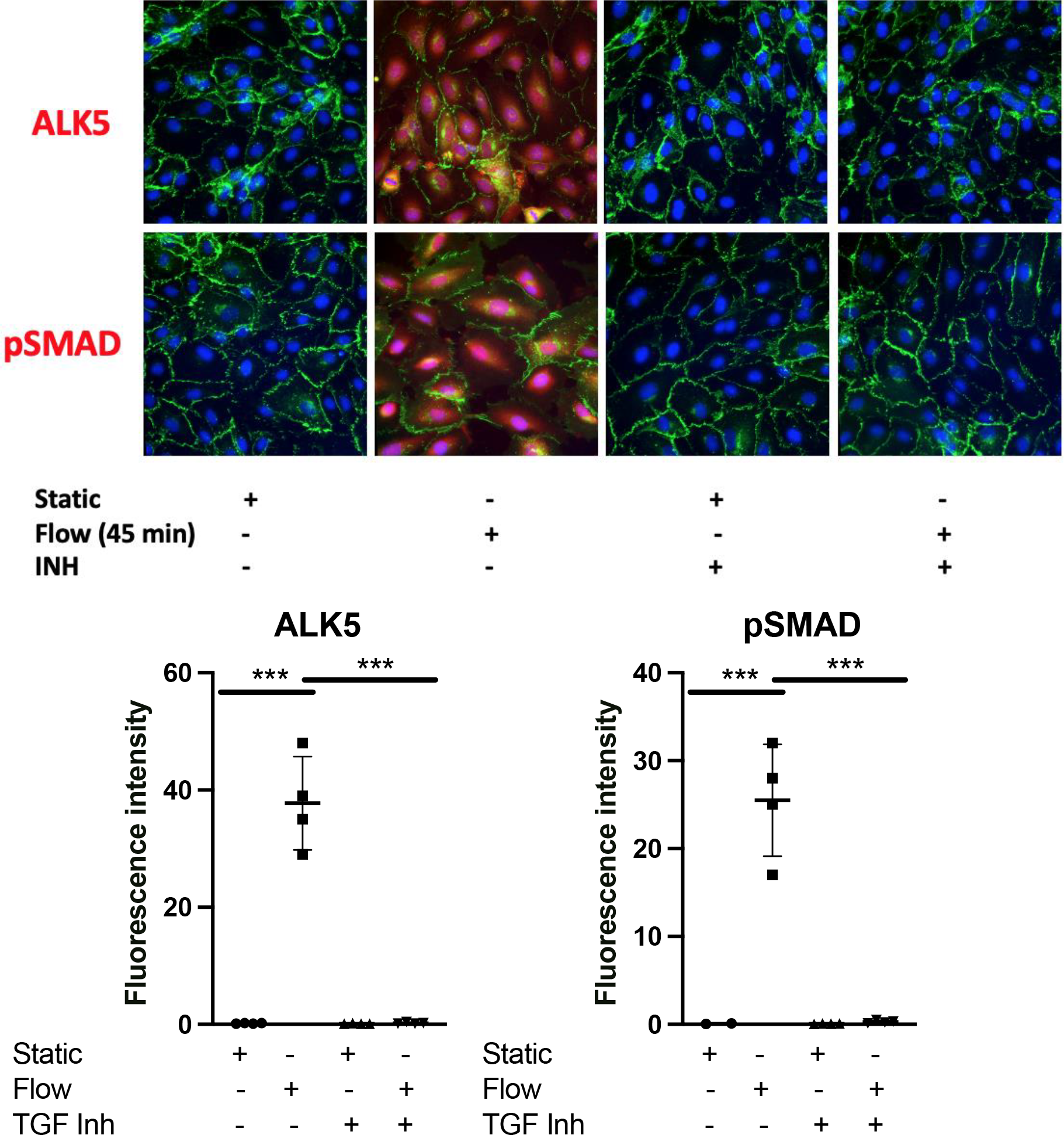
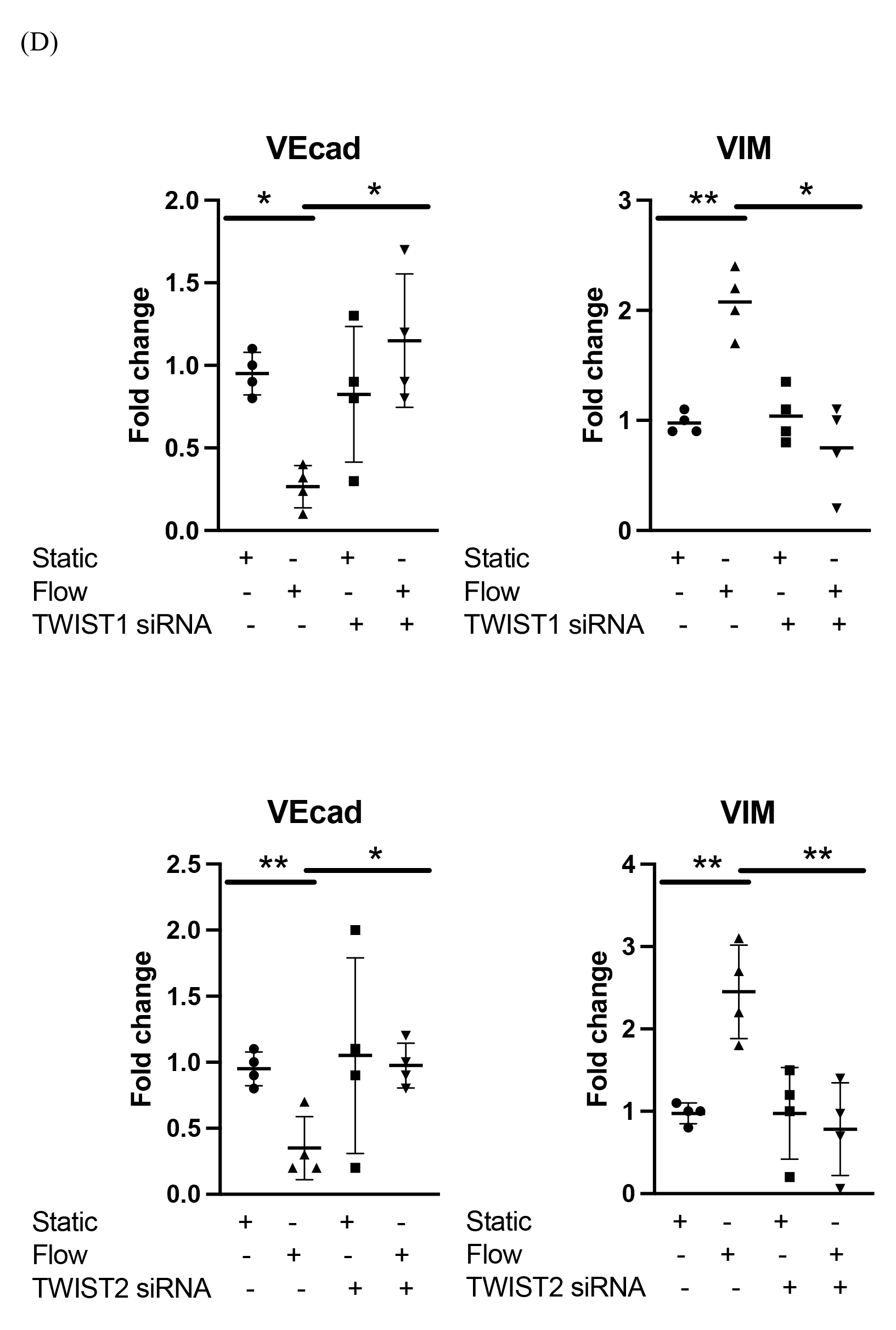
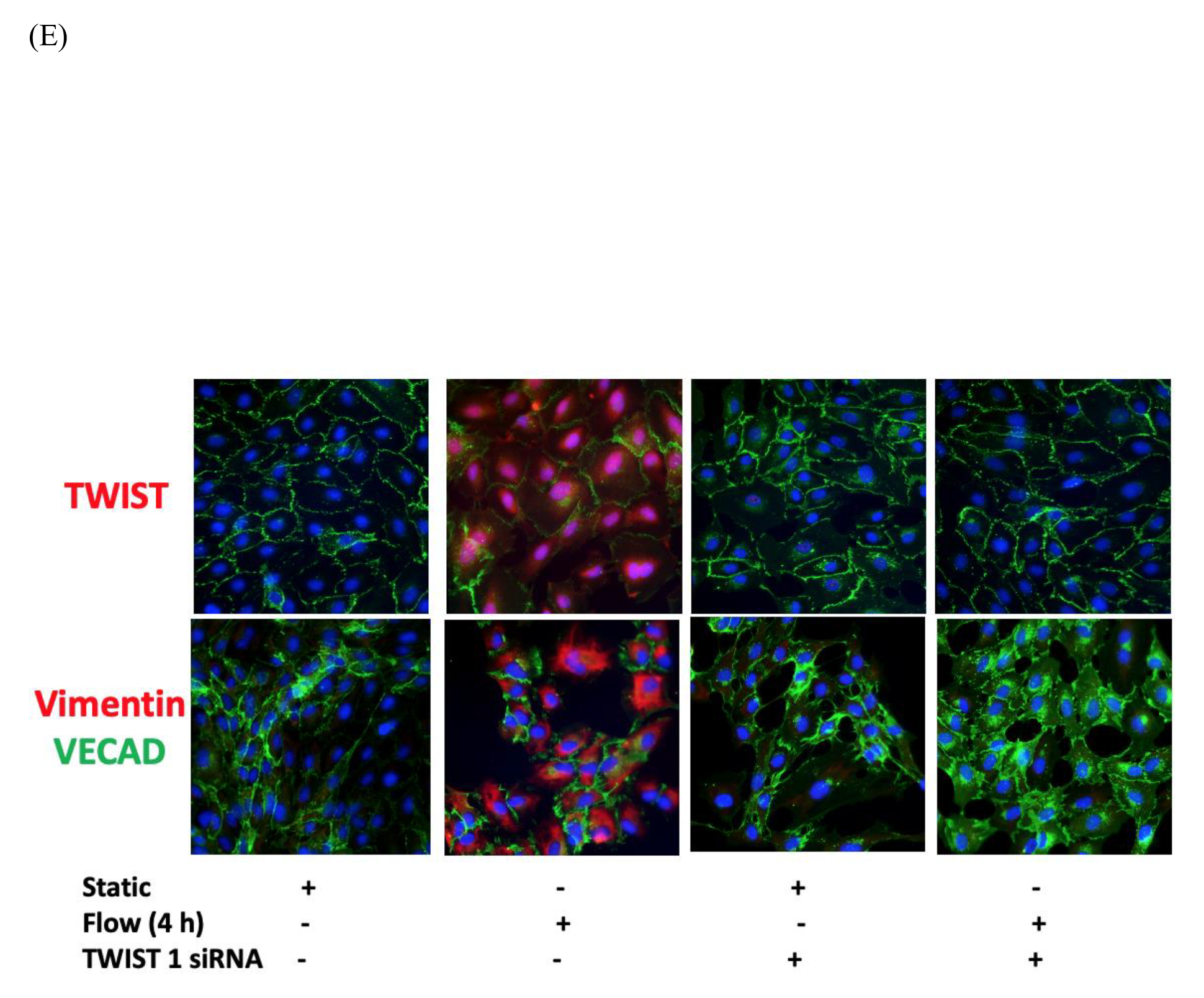
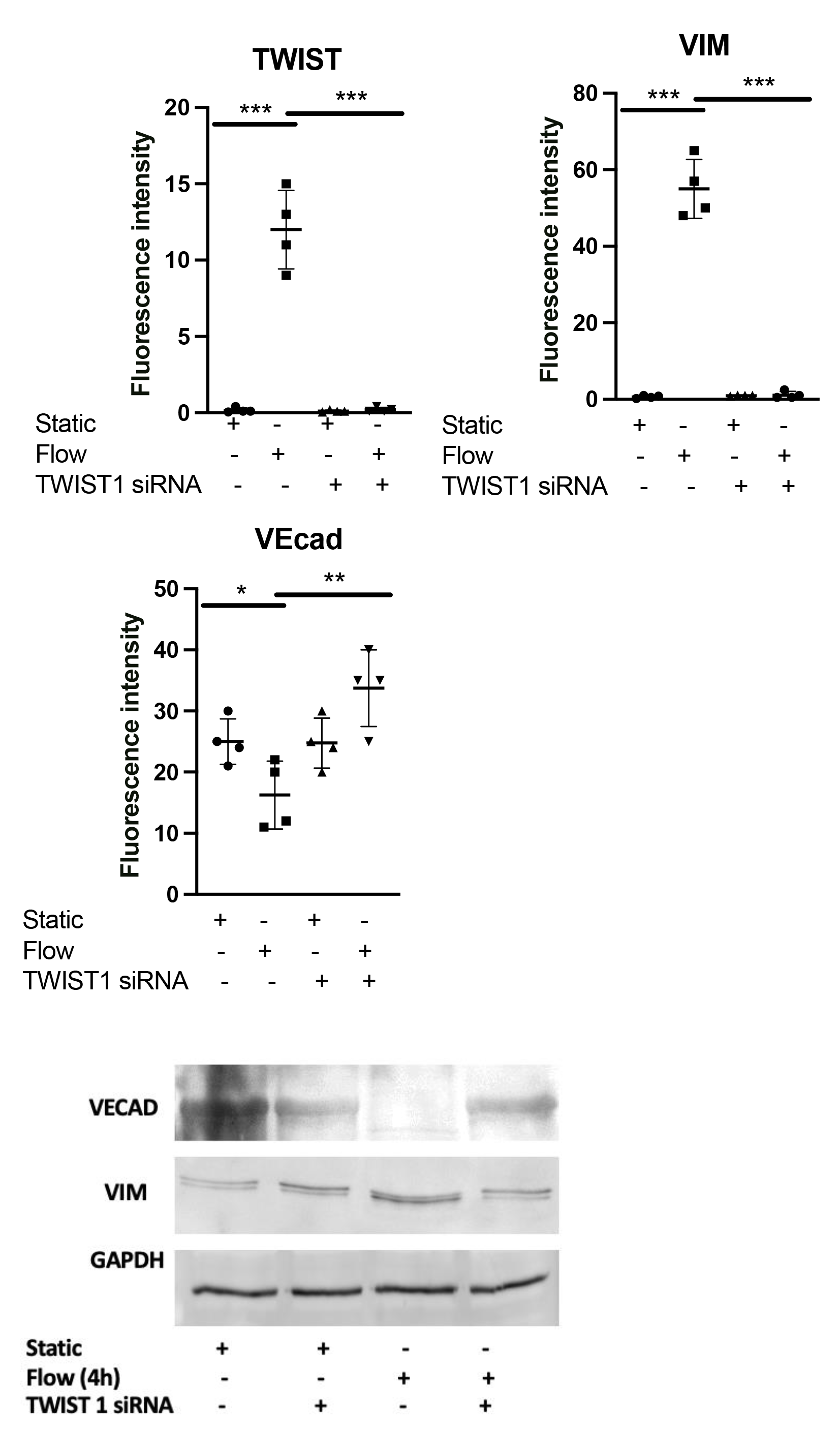
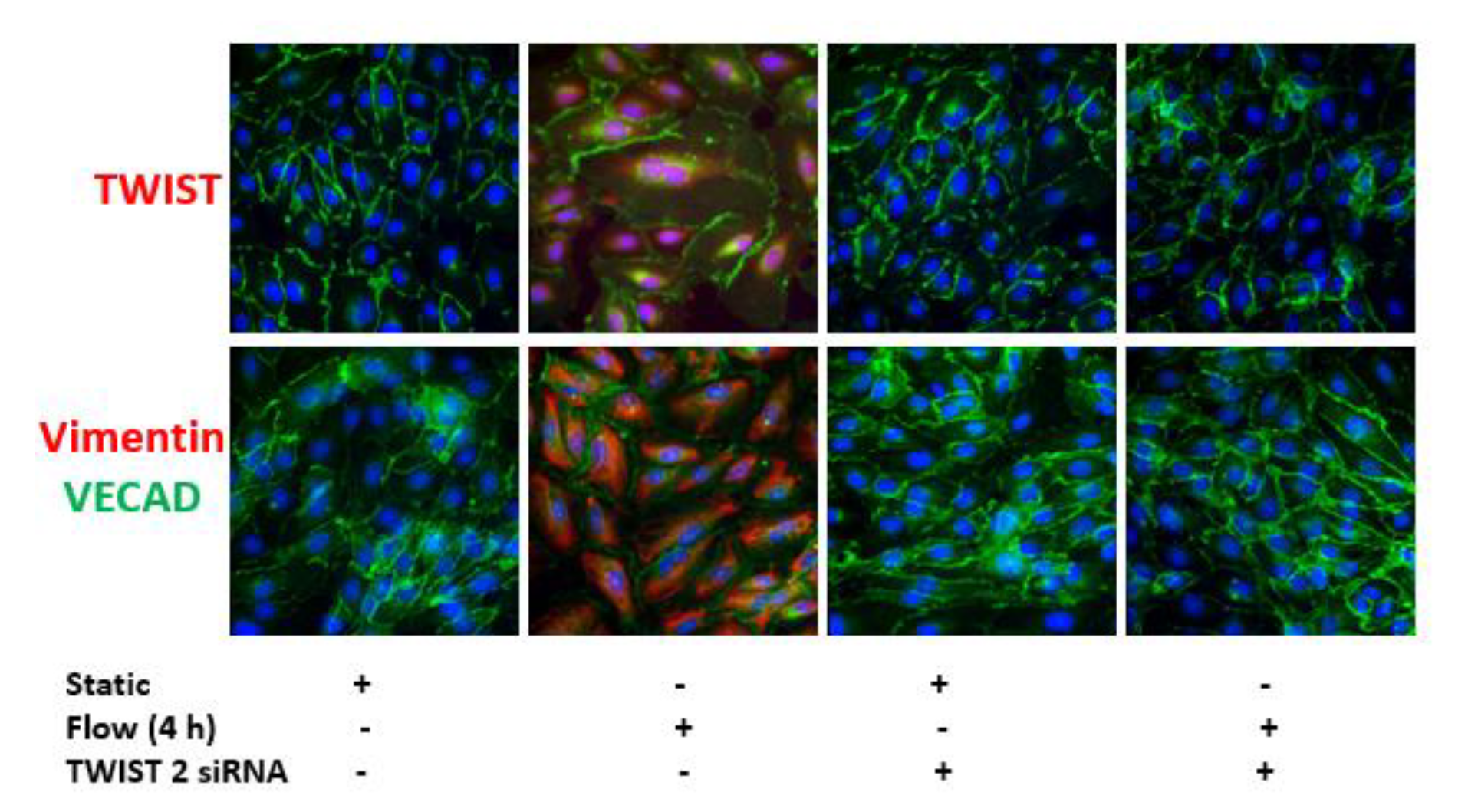
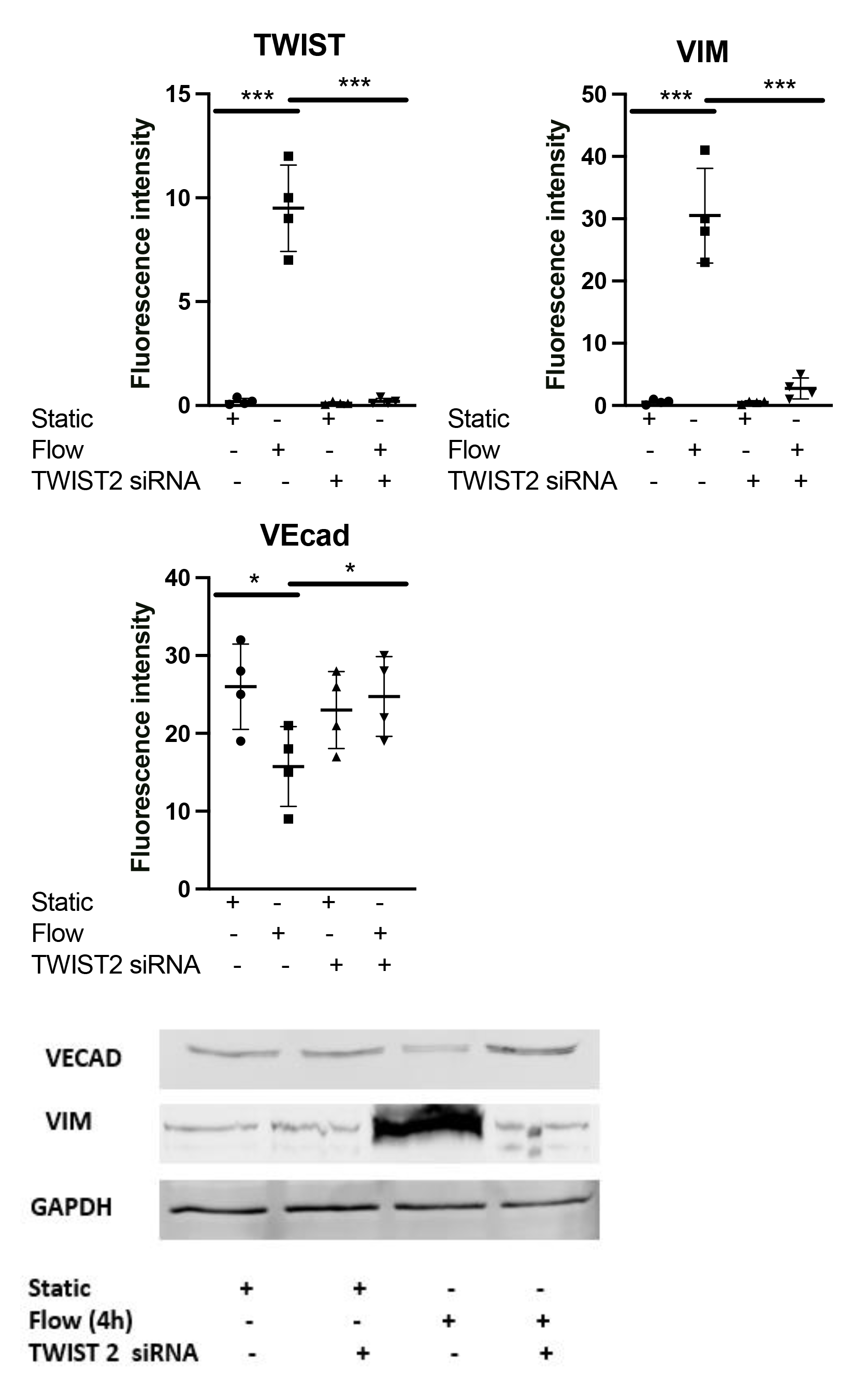
Effects of arterial shear stress on proinflammatory and EndMT genes in venous EC. (A) HUVECs were cultured under static conditions or exposed to LSS for 4 hours. Transcript levels of MCP1, CXCL8, VEcad, VIM, TWIST1 and TWIST2 were measured by comparative RT-PCR. Values from four independent experiments and mean values are shown. (B) HUVECs were pretreated with an ALK5 inhibitor (5 μmol/L) or with vehicle alone for xx minutes and were then exposed to LSS for 45 min or 4 hours or cultured under static conditions. Transcript levels of VEcad, VIM, TWIST1 and TWIST2 were measured by comparative RT-PCR. Values from four independent experiments and mean values are shown. (C) HUVECs were pretreated with an ALK5 inhibitor or with vehicle alone for 60 minutes and were then exposed to LSS for 4 hours or cultured under static conditions. TWIST1/2, VIM and VEcad expression levels were assessed at 4 hours and ALK5 and Psmad 2/3 assessed at 45 minutes by immunofluorescence staining with specific antibodies, quantified in multiple ECs, and averaged for each experimental group. Representative images, values from four independent experiments, and mean values are shown. (D) HUVECs were transfected with TIWST1 or TWIST2–specific siRNA or with a scrambled control (Scr). Cells were then exposed to LSS or cultured under static conditions for 4 hours. Transcript levels of VEcad, VIM, TWIST1 and TWIST2 were measured by comparative RT-PCR. Values from four independent experiments and mean values are shown. (E) TWIST1/2, VIM and VEcad expression levels were assessed at 4 hours in HUVECs transfected with TIWST1 or TWIST2–specific siRNA or with a scrambled control (Scr) and exposed to LSS or cultured under static conditions for 4 hours by immunofluorescence staining with specific antibodies (values from four independent experiments and mean values are shown) or Western blotting of total cell lysates with specific antibodies (images shown are representative of four independent experiments. See supp data for full gels).

Next, the role of TWIST 1&2 was assessed separately with specific siRNA that suppressed messenger RNA expression in sheared HUVECs, whereas a nontargeting scrambled control had no effect (Supp Figure. 1B). We observed, using RT-PCR that silencing of TWIST 1 or TWIST 2 suppressed EndMT changes HUVECs under ASS at transcript levels (Figure. 1D) and protein levels using immunostaining and WB (Figure. 1E) but did not affect the activation of upstream ALK5 activation of SMAD 2/3 phosphorylation (Supp Figure. 1C) suggesting that they both play TWIST 1 and TWIST 2 can play an essential role in EndMT process under ASS.

### Dexamethasone Pretreatment Suppressed EndMT in Venous ECs

We examined the effects of using Dex, a synthetic glucocorticoid that is known to modulate EndMT in multiple cell types. Based on our previous work, we used a pretreatment dose of 10 μmol/L. A brief pretreatment with Dex significantly altered HUVECs responses to ASS by suppressing ALK5 activation and SMAD2/3 phosphorylation at 45 min (Figure. 2A). Moreover, it was associated with the suppression of TWIST 1, TWIST 2 and VIM while restoring the expression of VEcad both at the messenger RNA (Figure. 2B) and protein levels using immunostaining as WB (Figure. 2C). These results suggests that the use of Dex can suppress EndMT changes in response to ASS in addition to previously known effect of suppressing MAPK mediated proinflammatory activation in HUVECs in response to ASS making it a potential therapeutic that is capable of modulating different adverse effects of ASS on venous EC.

**Figure. 2.**
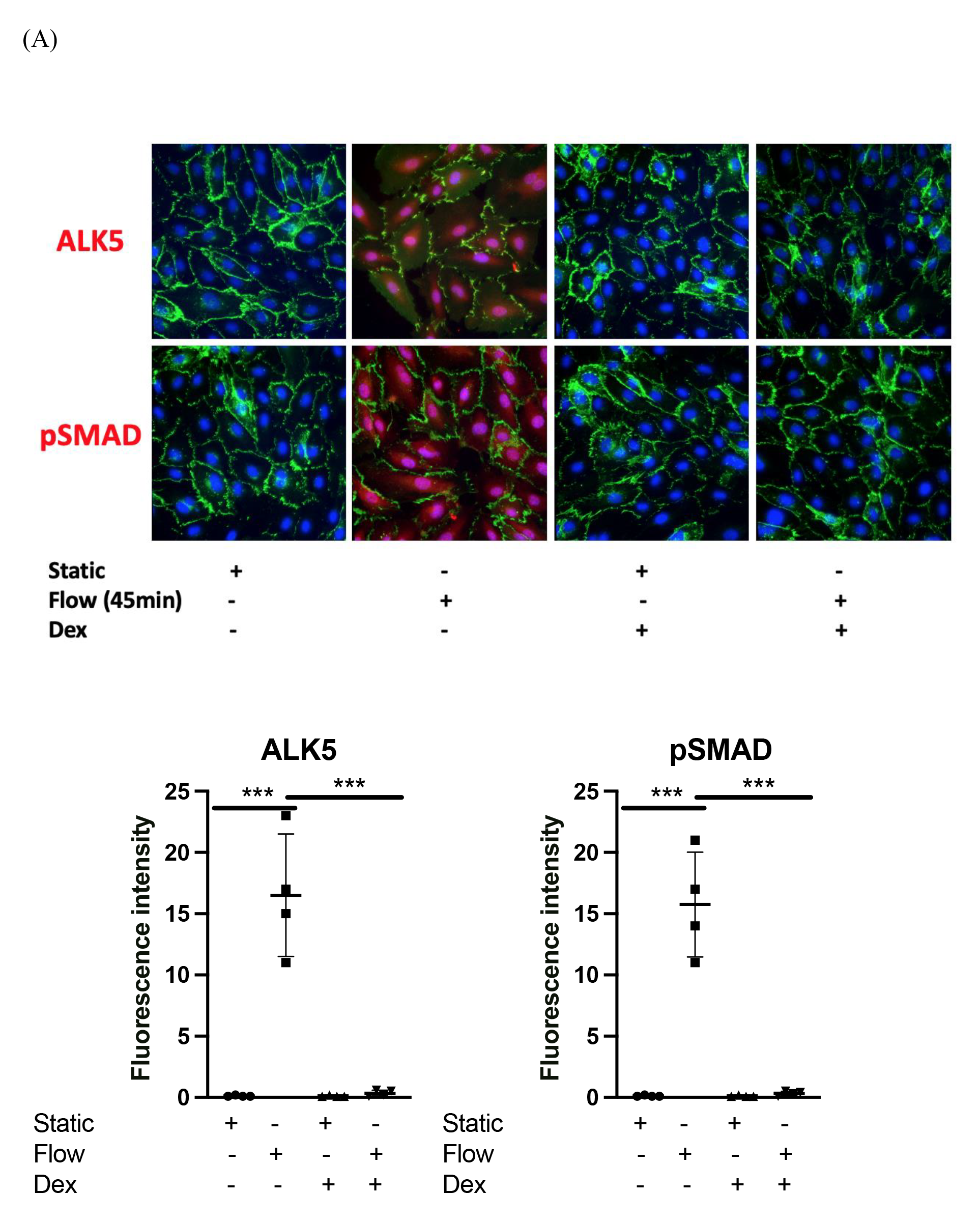
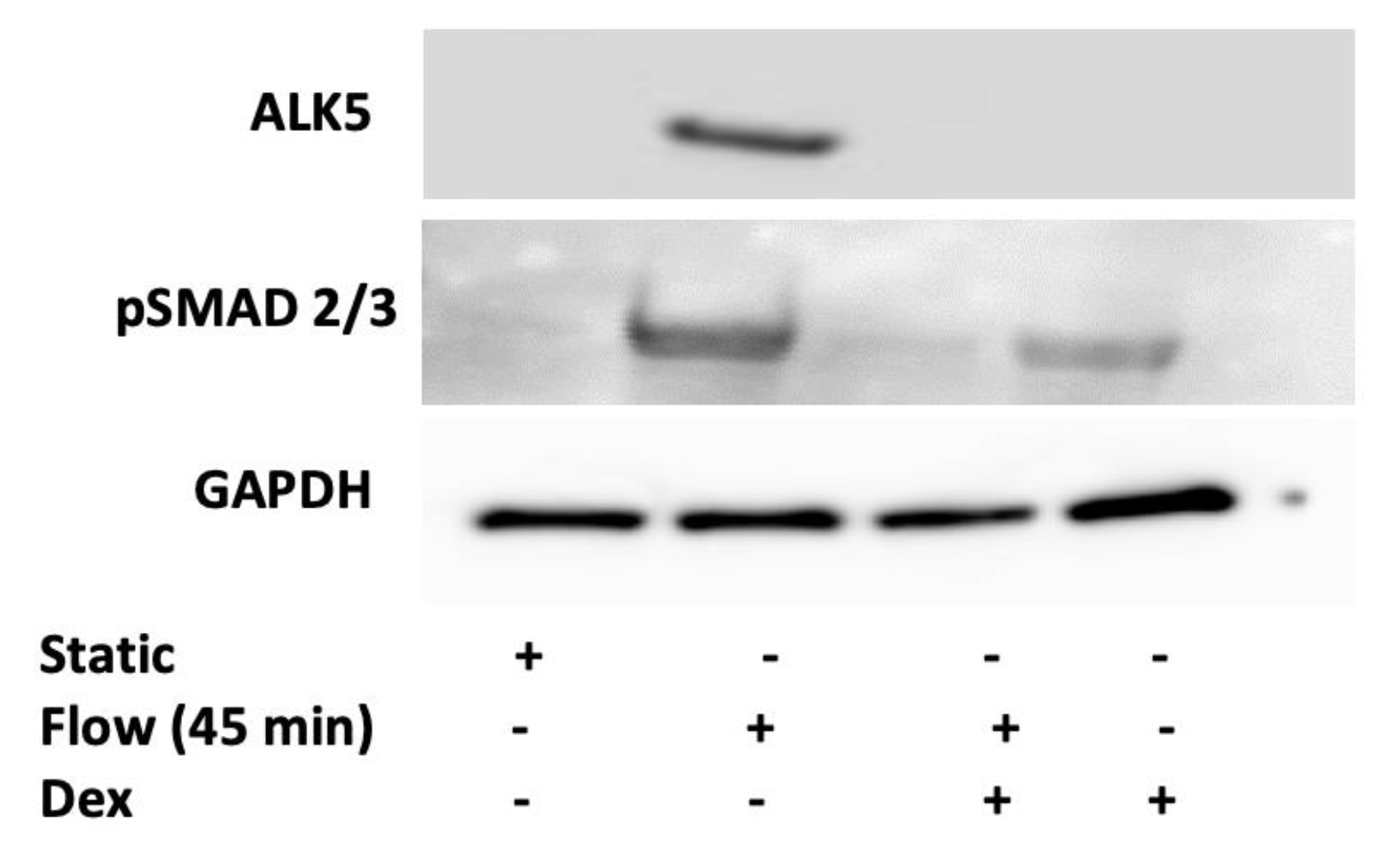
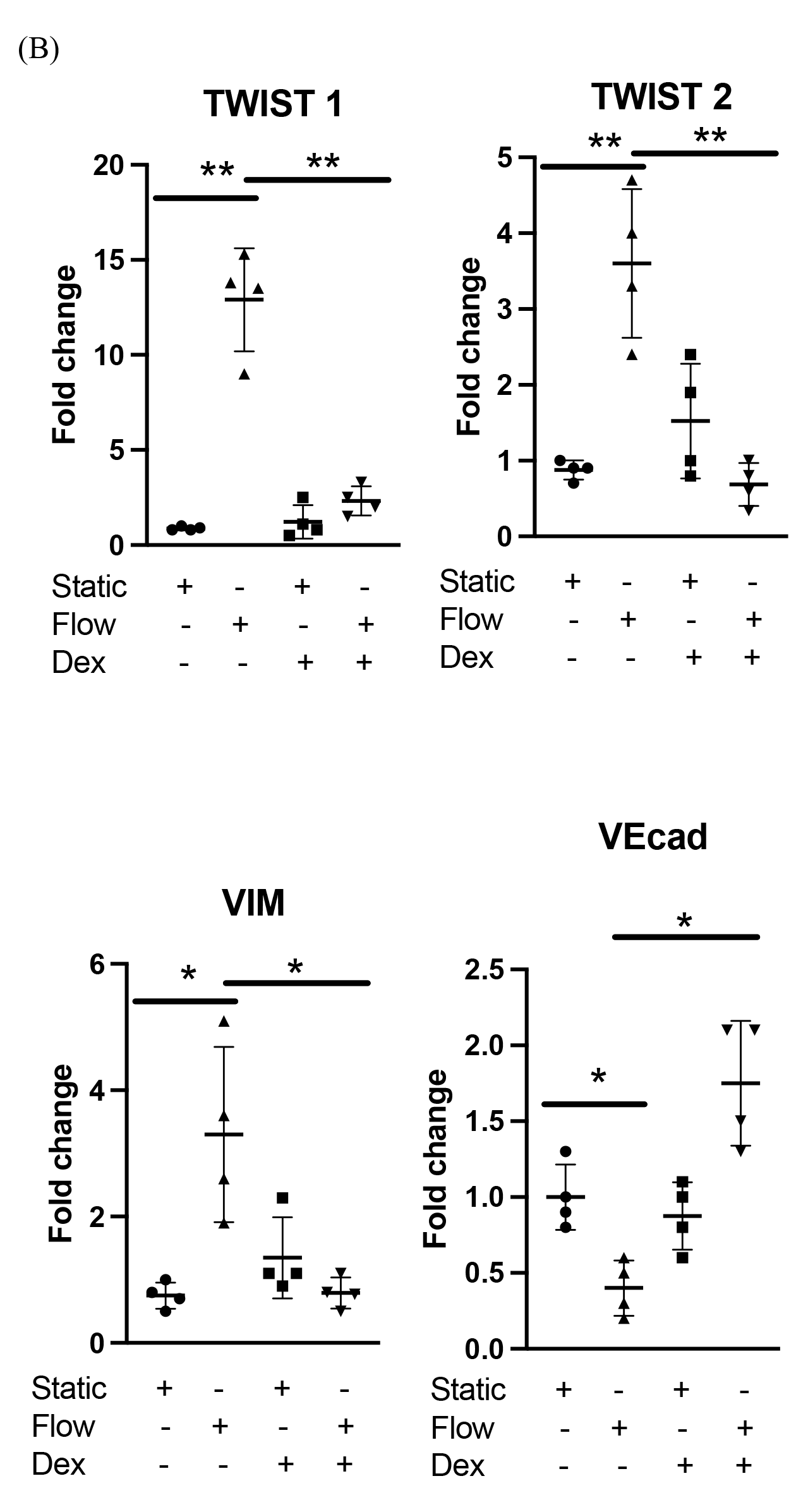
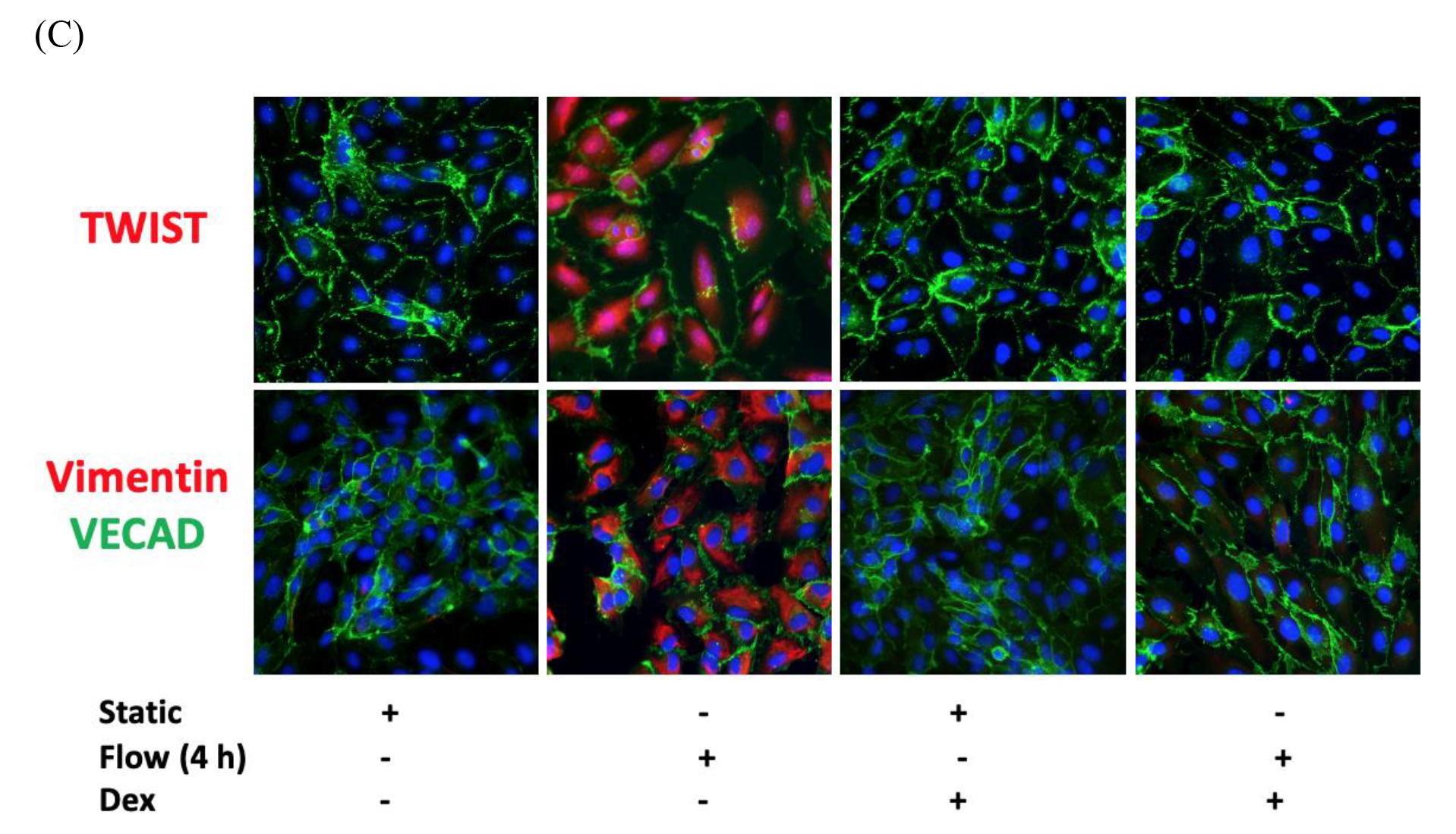
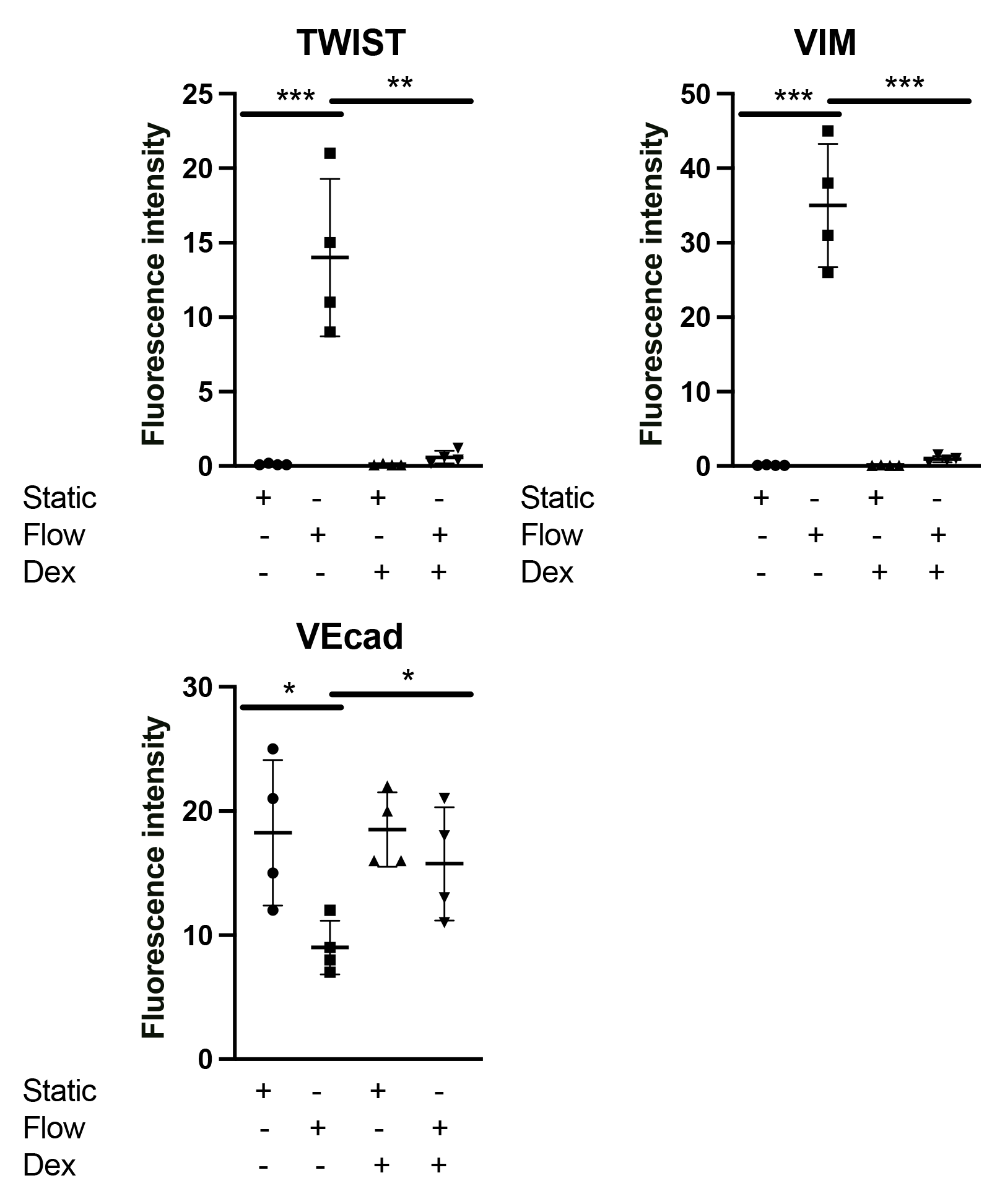
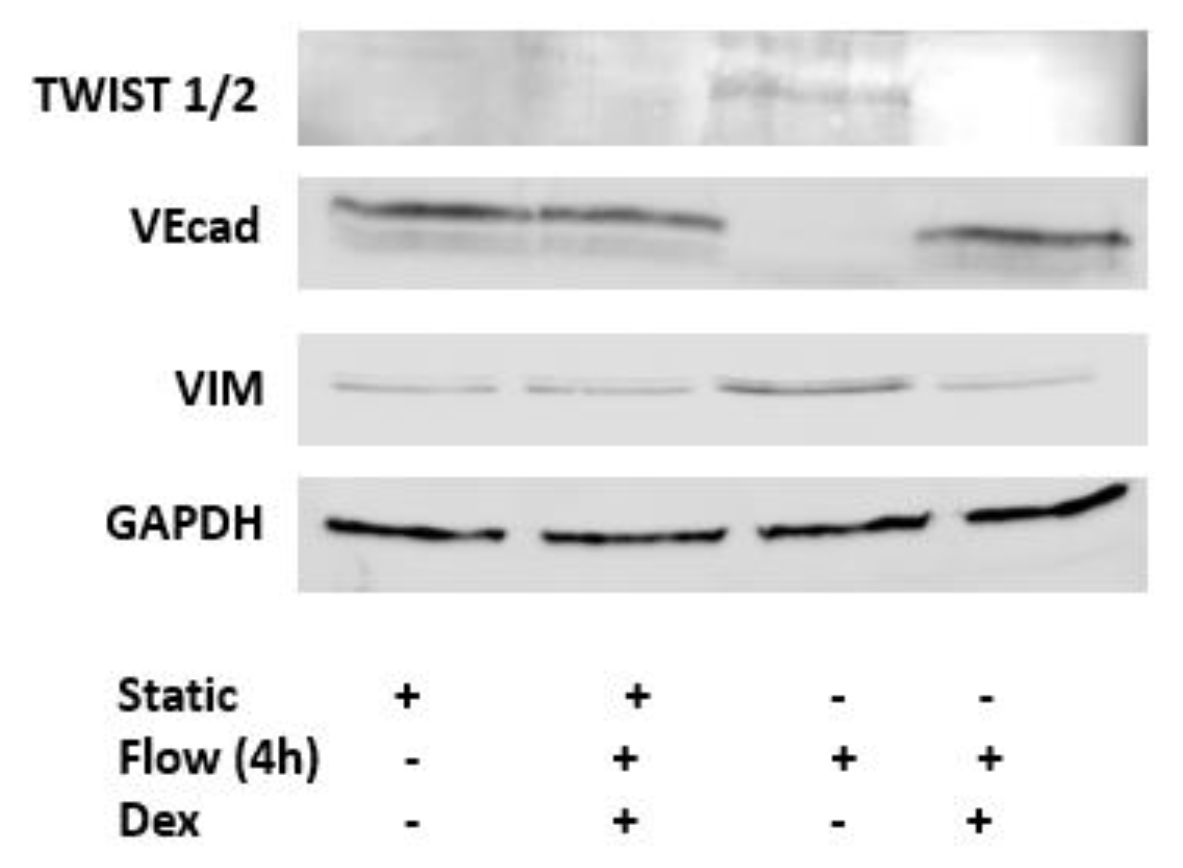
Dexamethasone suppressed EndMT activation by shear stress in cultured venous ECs. (A) HUVECs were pretreated with dexamethasone (10 μmol/L) for 30 minutes or remained untreated and were then exposed to LSS for 45 minutes. ALK5 and Psmad 2/3 was assessed by immunofluorescence staining with specific antibodies (values from four independent experiments and mean values are shown) or Western blotting of total cell lysates with specific antibodies (images shown are representative of four independent experiments. See supp data for full gels). (B) HUVECs were pretreated with dexamethasone (10 μmol/L) for 30 minutes or remained untreated and were then exposed to LSS for 4 hours. Transcript levels of TWIST1, TWIST2, VEcad and VIM were measured by comparative RT-PCR. Values from four independent experiments and mean values are shown. (C) TWIST1/2, VIM and VEcad expression levels were assessed at 4 hours in HUVECs that were pretreated with dexamethasone (10 μmol/L) for 30 minutes or remained untreated by immunofluorescence staining with specific antibodies (values from four independent experiments and mean values are shown) or Western blotting of total cell lysates with specific antibodies (images shown are representative of four independent experiments. See supp data for full gels).

### EndMT activation in veins exposed to arterial shear stress *ex-vivo* using untargeted spatial cell sequencing

We validated our *in-vitro* findings by studying freshly harvested human saphenous veins exposed to short-term arterial flow *ex-vivo*. In a series of first-in-man experiments (n=4), we performed spatial cell RNA sequencing on vein segments exposed to acute arterial haemodynamics for 4 hours (Supp Figure. 2A, B, C). We carried out clustering of cells based on multiple common markers for EC, SMC, and fibroblasts, in combination with analysis of the top 30 differentially expressed genes in each cluster (Figure 3A). In this clustering we identified a cluster of cells that has both EC and SMC phenotypes (EC/SMC) (Figure. 3B, C) suggesting that it may represent an intermediate EndMT as it has been reported previously. (21) In this cluster, 16954 genes were identified out of these 519 (3.1%) were significantly differentially regulated under flow conditions (Figure. 3D). Global pathway analysis showed correlations made to the different groupings including the regulation of signalling, specifically the inflammatory and immune responses (cytokine production and responses), leukocyte activation, migration, differentiation, and adhesion, wound healing responses, and activation/regulation of the p53-MAPK signalling cascade. Additionally, there was correlation to the regulation of cell behaviour, including activation, proliferation, differentiation, migration, cell-cell communication, adhesion, and motility, with specific reference to smooth muscle cell proliferation and endothelial cell migration. Correlation was also present to responses to stress, oxidative stress (and reactive oxygen species), integrated stress response signalling and apoptosis (Figure. 3E). Global network analysis was able to identify a total of 2204 connections between 436 of the 519 differentially expressed genes in the EC/SMC cluster (Supp Figure. 2D). When focusing on TWIST1 and TWIST 2 network analysis in this cluster specifically, we noted associations with genes such as TGIF1, FOXC2, CD44, CYP1B1, NFIC, REL and FOXC1and their subsequent associations. Such genes are known to be involved in processes such as inflammation, EndMT, immune signalling, differentiation and apoptosis. (29-36) (Figure. 3F)

**Figure. 3.**
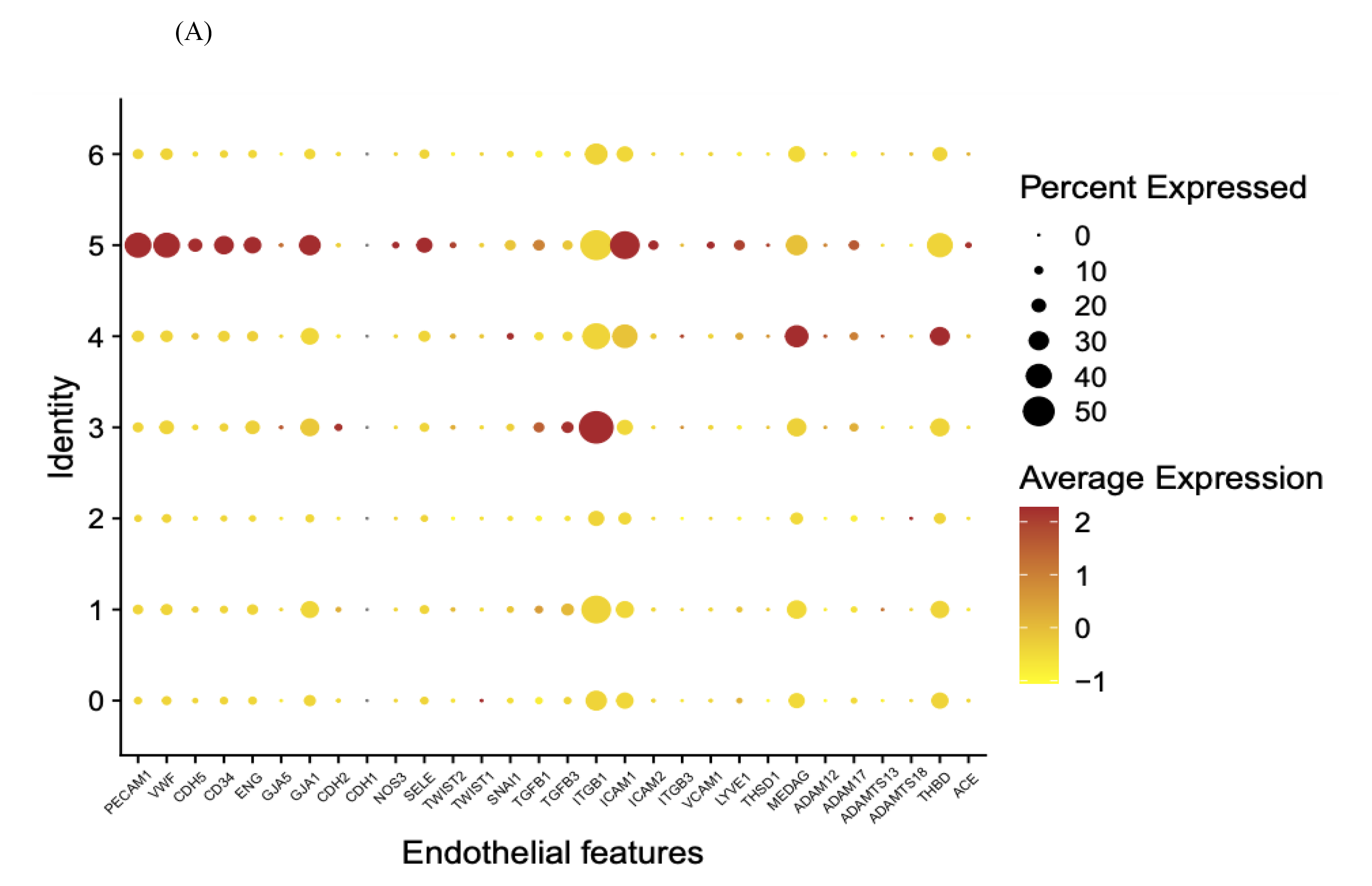
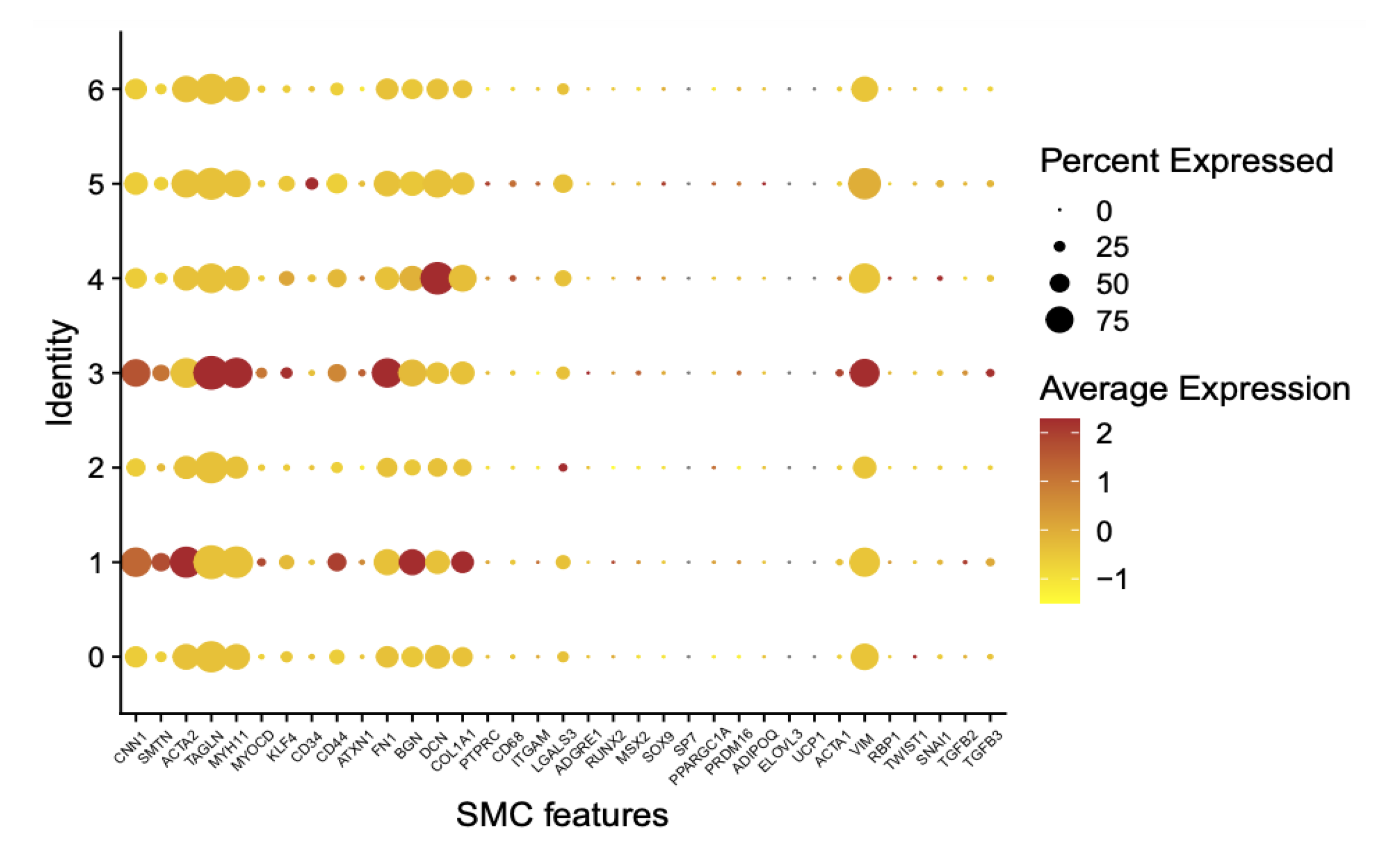
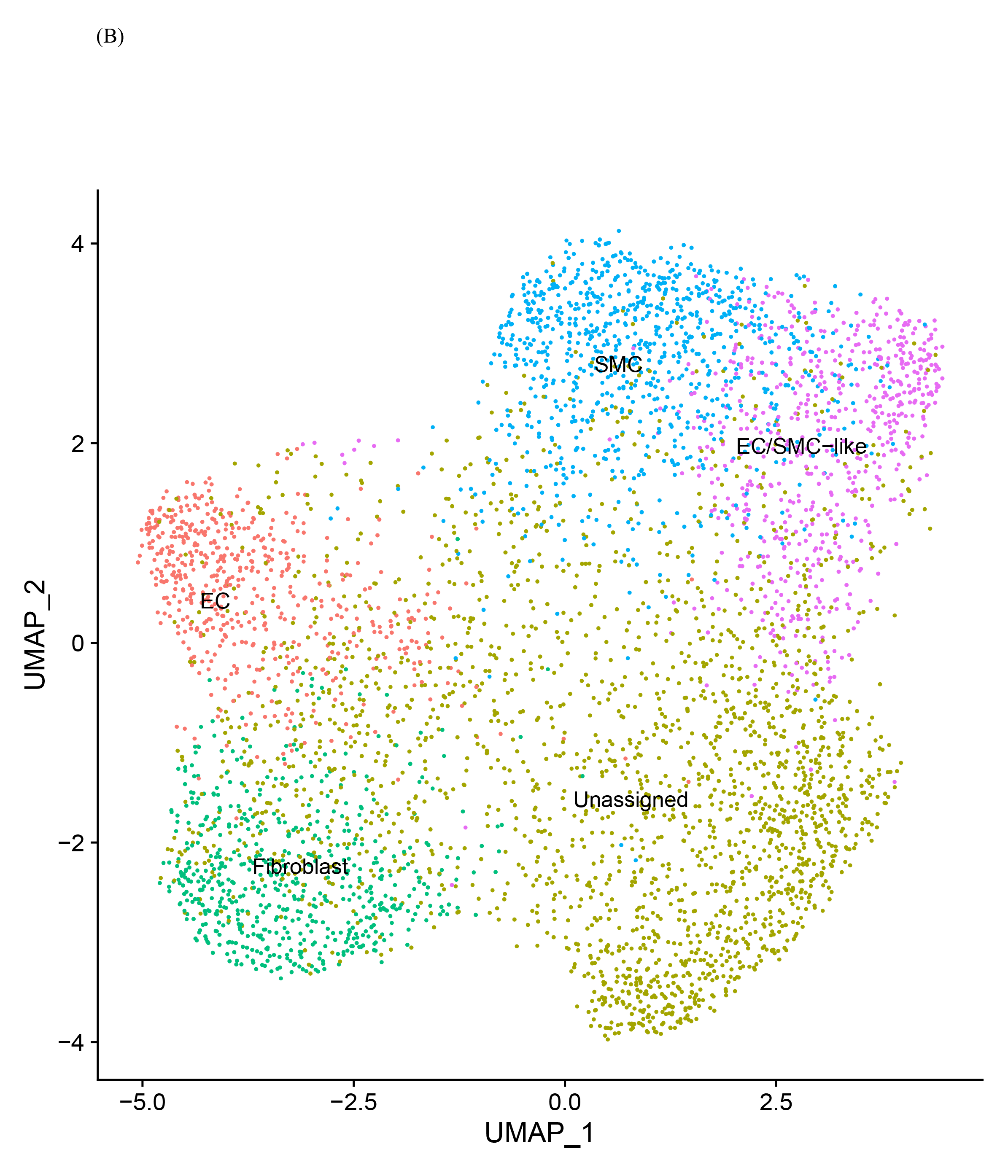
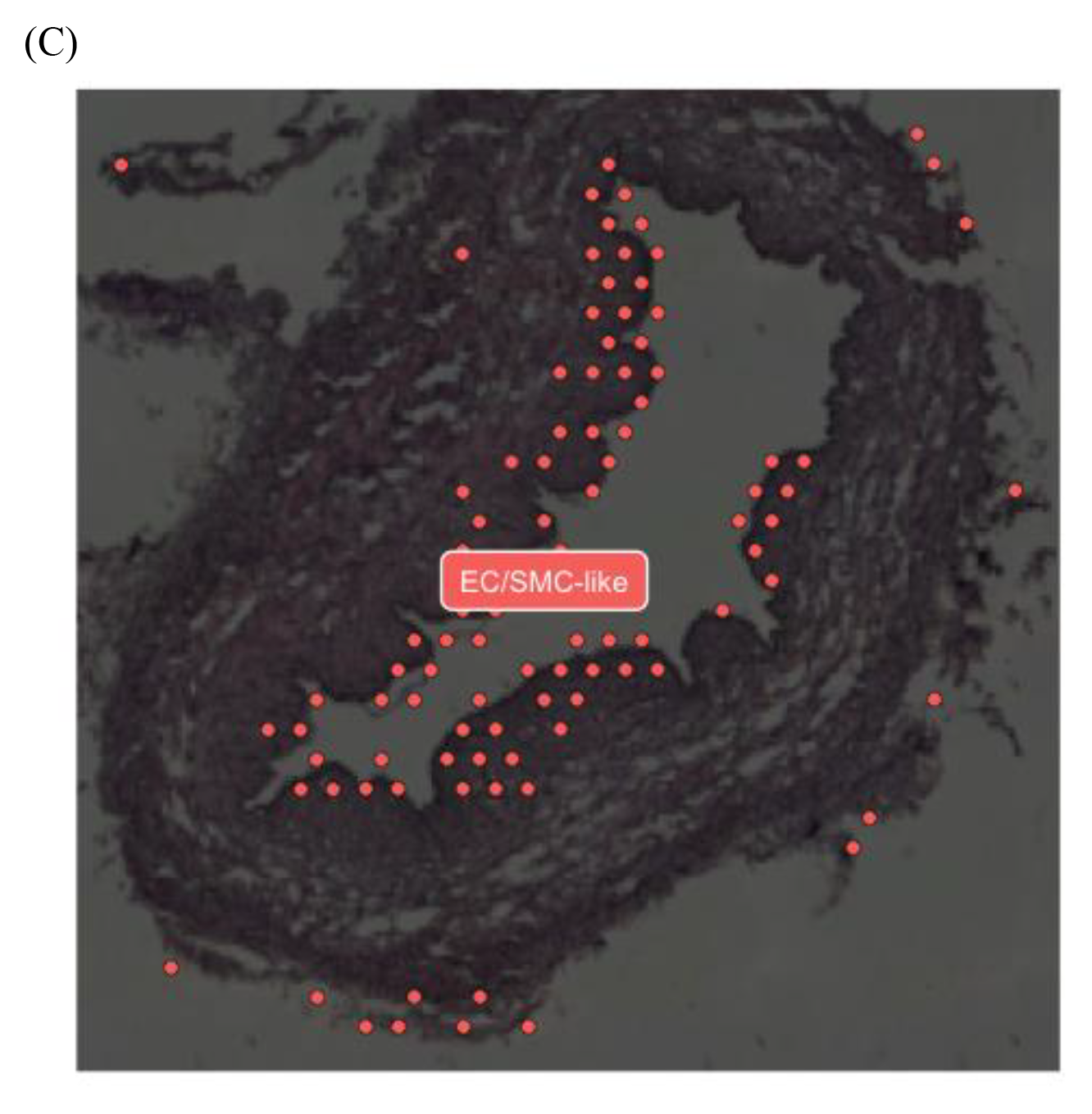
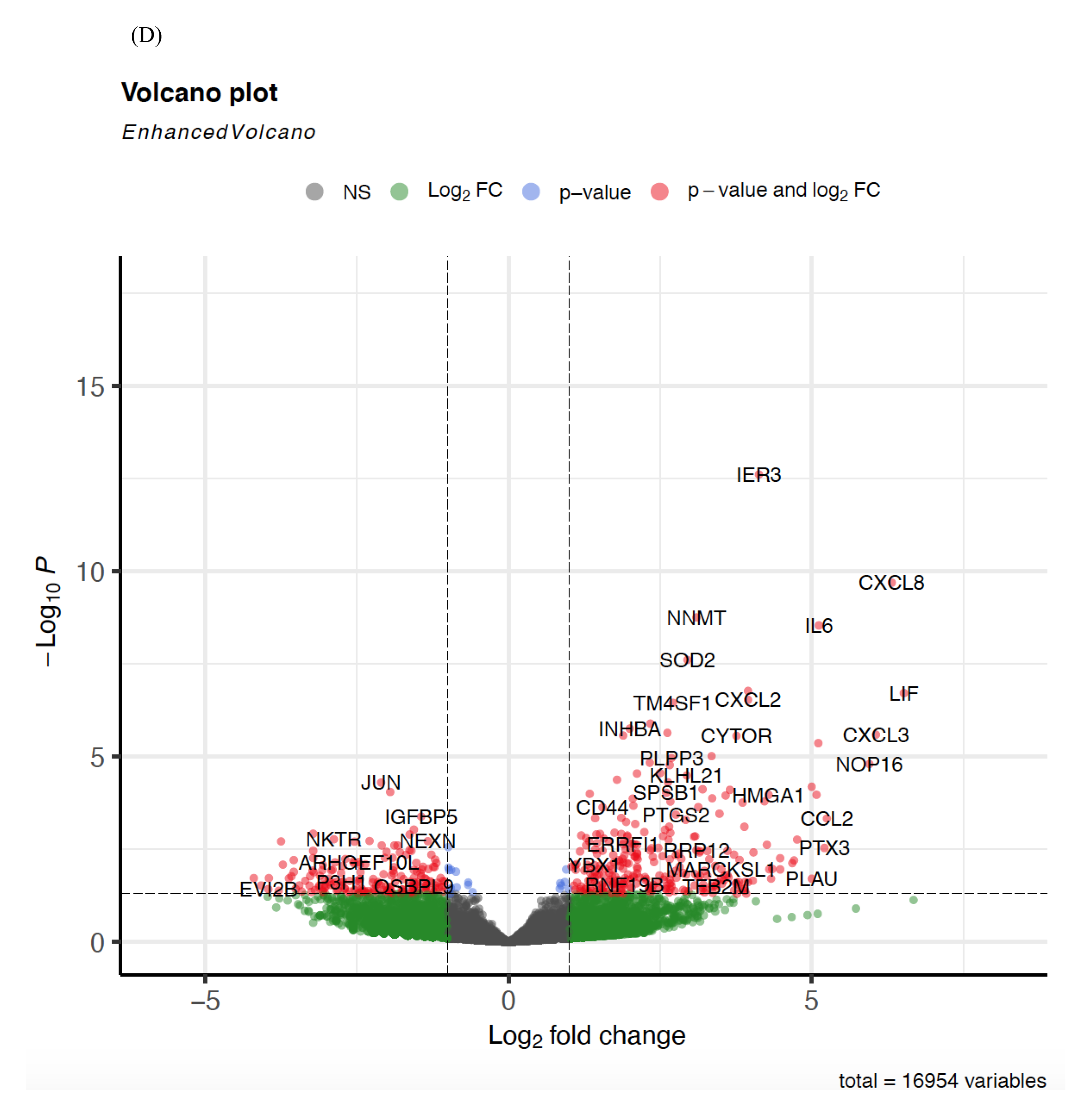
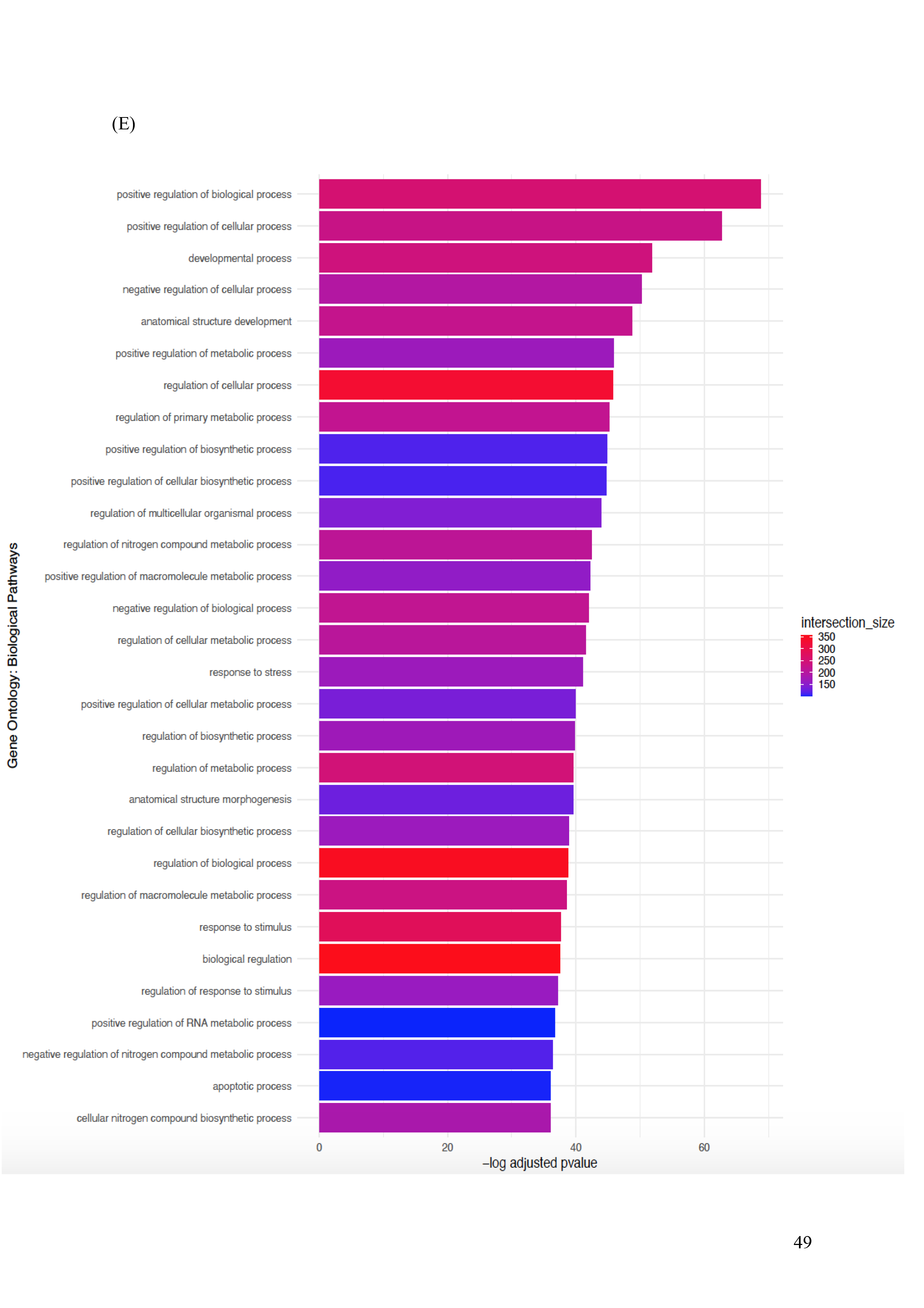
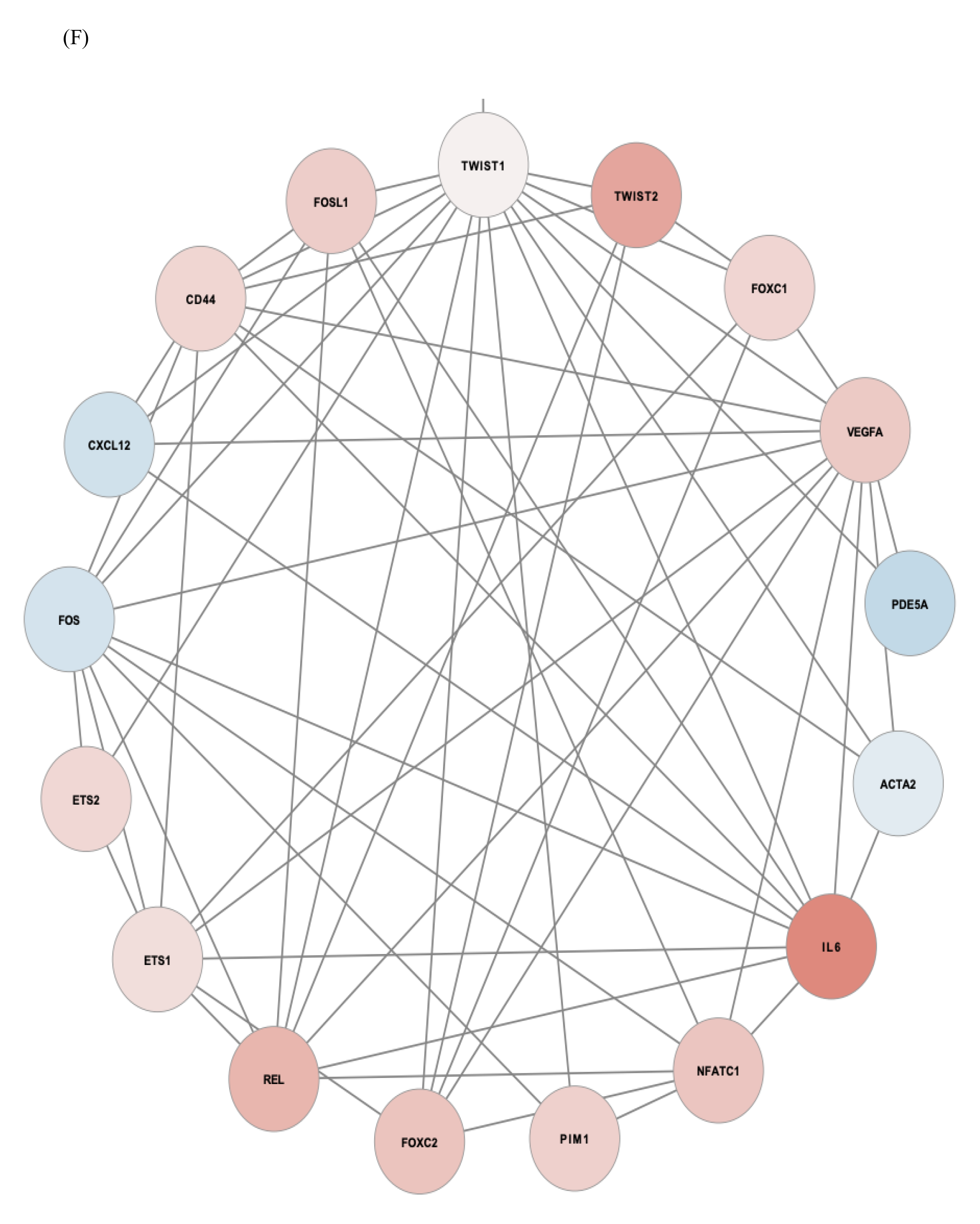
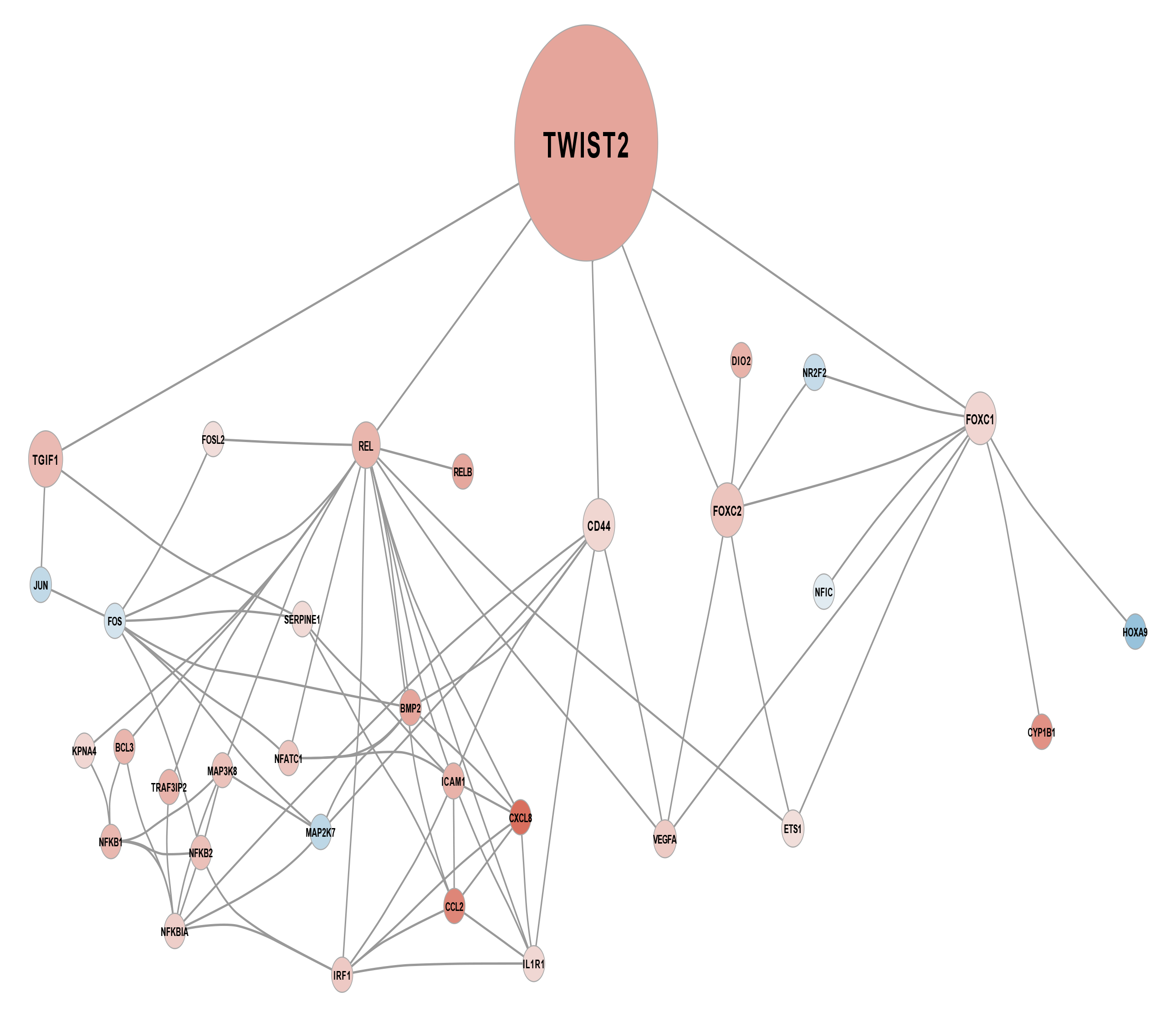
EndMT activation in veins exposed to arterial shear stress ex-vivo using untargeted spatial cell sequencing. (A) Clustering of cells based on multiple common markers for EC, SMC, and fibroblasts, in combination with analysis of the top 30 differentially expressed genes in each cluster. (B) Cluster of cells that has both EC and SMC phenotypes (EC/SMC). (C) representative images of LSV tissue section (10um) microscopy image and its associated 10X Visium spot coverage grid overlay showing the spatial distribution of the EC/SMC cluster. (D) Volcano plot of the clustered identified as EC/SMC depicting all differentially expressed genes (DEGs) between the control samples and samples exposed to 4 hours arterial haemodynamics (n = 4). (E) Global pathway analysis of the EC/SMC cluster of all significant differentially expressed genes (adjusted p-value < 0.05) with the top 30 connections presented. Enrichment is scaled based on significance (-log10 p_value) and visually ranked based on intersection size (overlap of DEGs with enriched pathways). (F) EC/SMC cluster TWIST1 and TWIST2 subset network of significant differentially expressed genes (adjusted p-value < 0.05) in samples exposed to *ex vivo* arterial haemodynamic conditions. Direct connections illustrate known direct interactions, and these interactions are highlighted in the table.

### Dexamethasone Pretreatment Suppressed EndMT in Veins Exposed to Arterial Shear Stress *ex-vivo*

We next set up to validate the above finding in relation to genes related to EndMT using targeted approach including RT-PCR for the whole tissue and considering the limitation of spatial sequencing in term of single cell localisation, we utilised RNAscope that allow the localization of genes of interest expression in the vein sections and studied the impact of a brief pretreatment of veins with 10 μmol/L Dex (Other validated genes of interest in Supp Figure. 3A). In addition to suppressing proinflammatory response to acute arterial flow as noted by the suppression of CXCL8, MCP-1 and VCAM1 (Supp Figure. 3B), Dex modulated EC responses by suppressing TWIST1 and TWIST2 at transcript levels using RT-PCR for the whole tissue (Figure. 4A) and RNAscope at 4 hours which demonstrated the activation and localisation of both transcription’s RNA in the EC which was suppressed by Dex treatment (Figure. 4B). Additionally, this brief pretreatment was able to supress EndMT phenotype changes in EC for VEcad and VIM at RNA levels in whole tissue using RT-PCR (Figure. 4C). Moreover, Dex pretreatment significantly modulated TWIST1 and TWIST 2, VEcad and VIM protein levels using immunostaining (Figure. 4D). Furthermore, Immunostaining revealed that Dex action was also associated with a rapid (45 min) significant reduction in ALK5 expression and SMAD 2/3 activation (phosphorylation) (Figure. 4 E). Interestingly, when we looked at SNAIL1 transcription using RNAscope, we noted no activation in EC but only presence in SMC which was suppressed by Dex pretreatment (Supp Figure. 3C) suggesting the TWIST and SNAIL transcription factors may work on specific cell types. We next studied the role of Dex pretreatment on the regulation of monocytes adhesion using adhesion assay as previously reported and noted that the brief pretreatment with Dex can significantly reduce monocytes adhesion to LSV (Figure. 4F). Moreover, considering the limitation of running long time scale flow experiments, we compared the effect of brief Dex pretreatment followed by LSV culture for 10 days and noted that the brief single treatment was sufficient in suppressing EndMT at 10 days post culture (Supp Figure. 3D).

**Figure. 4.**
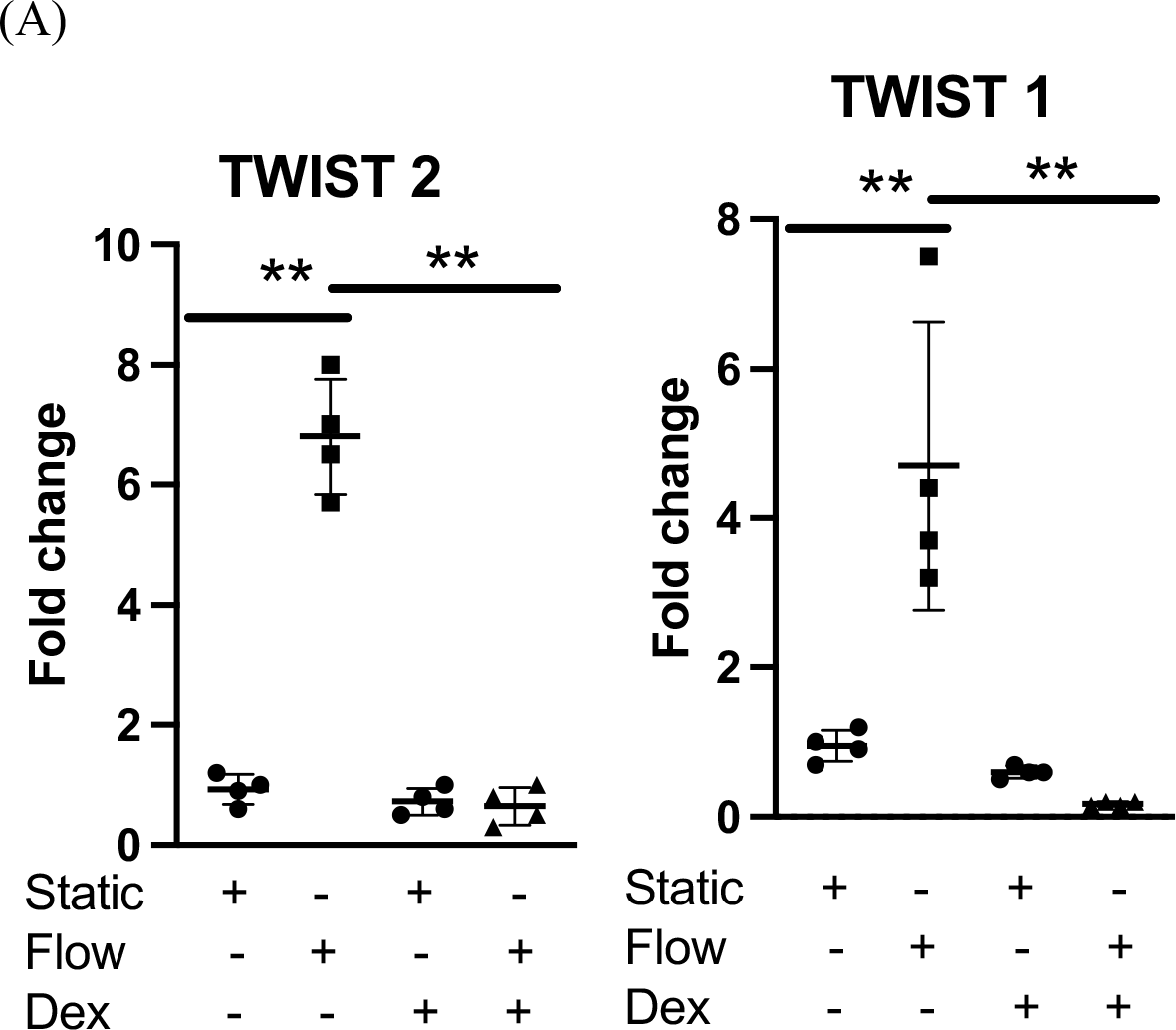
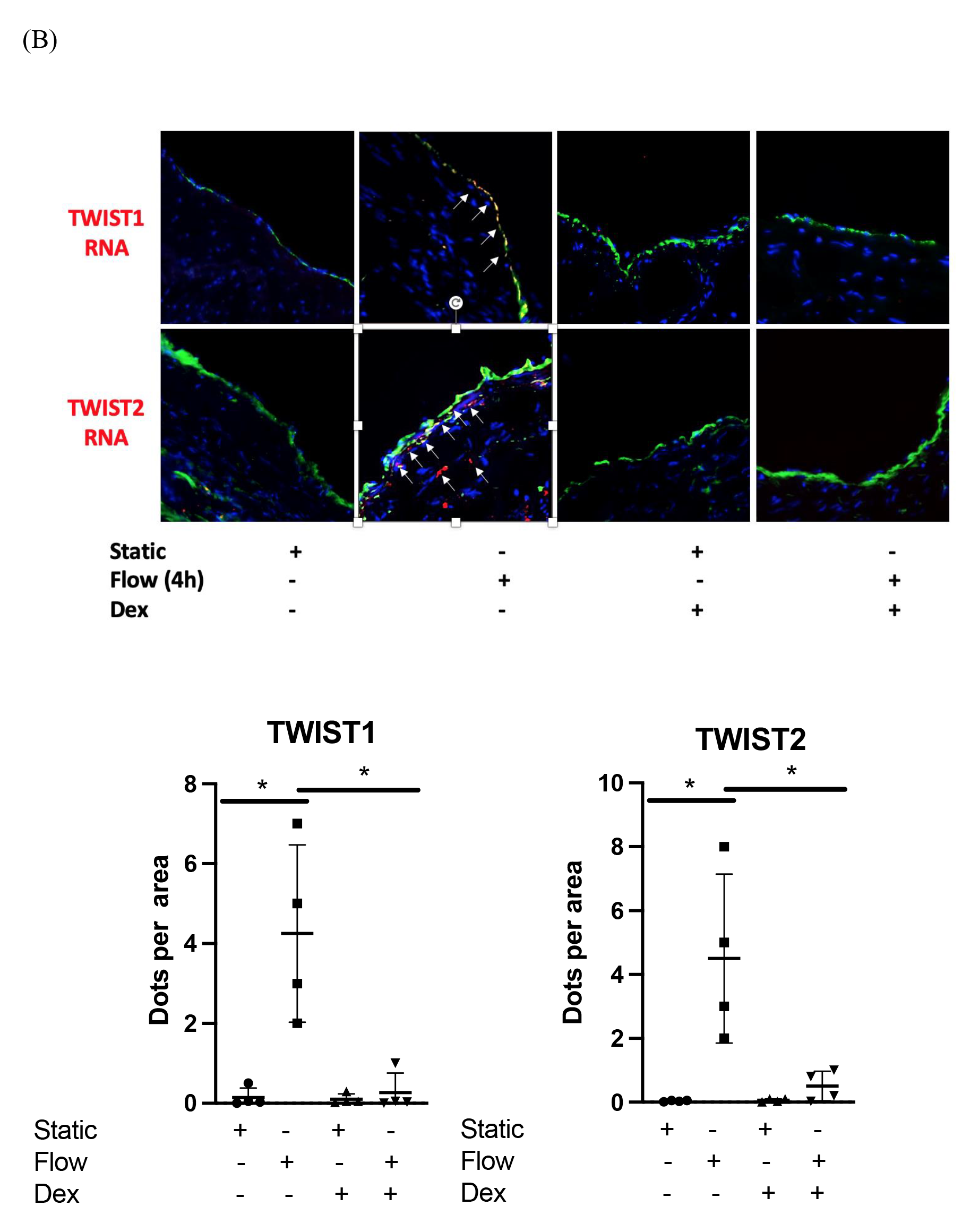
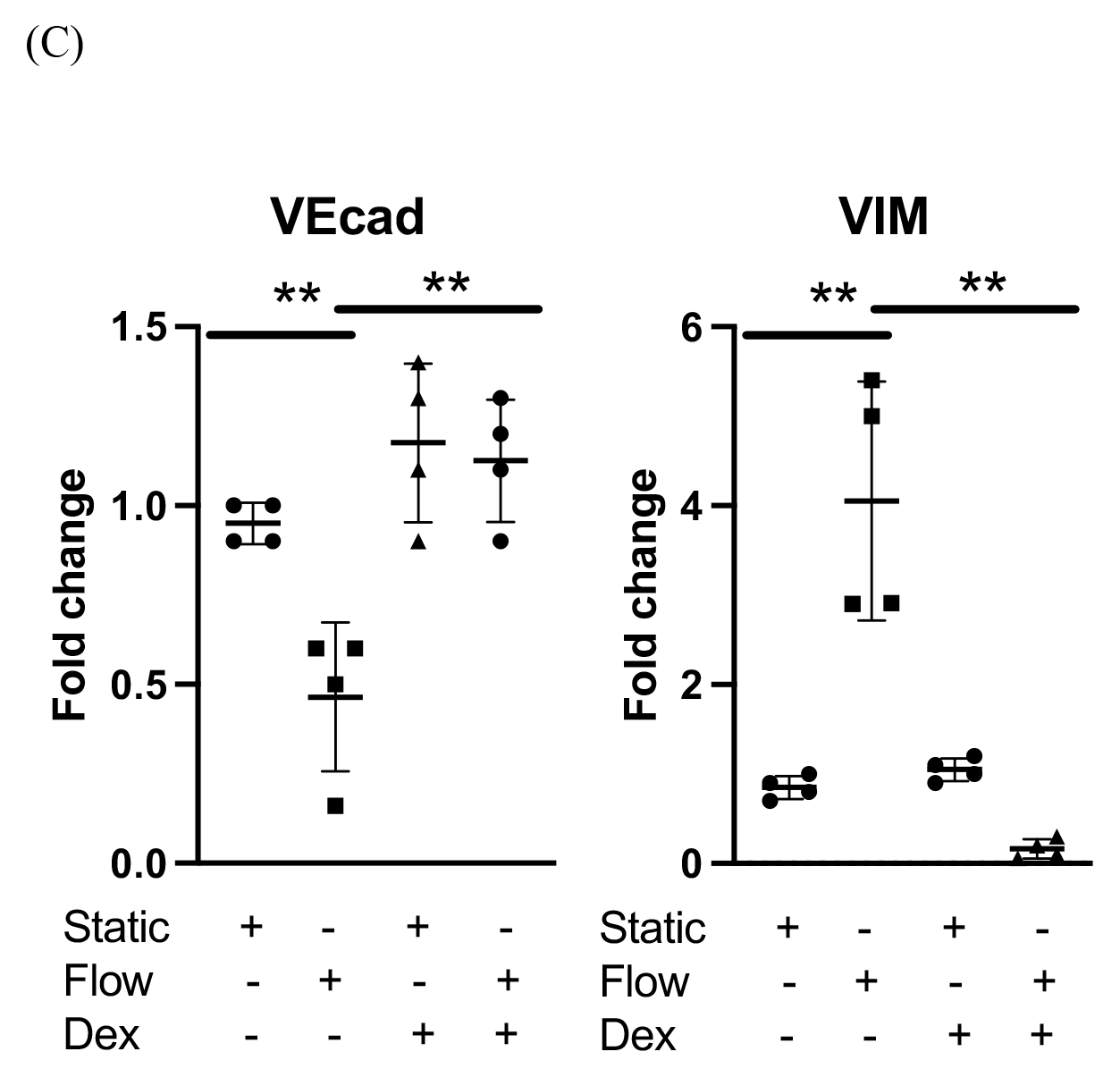
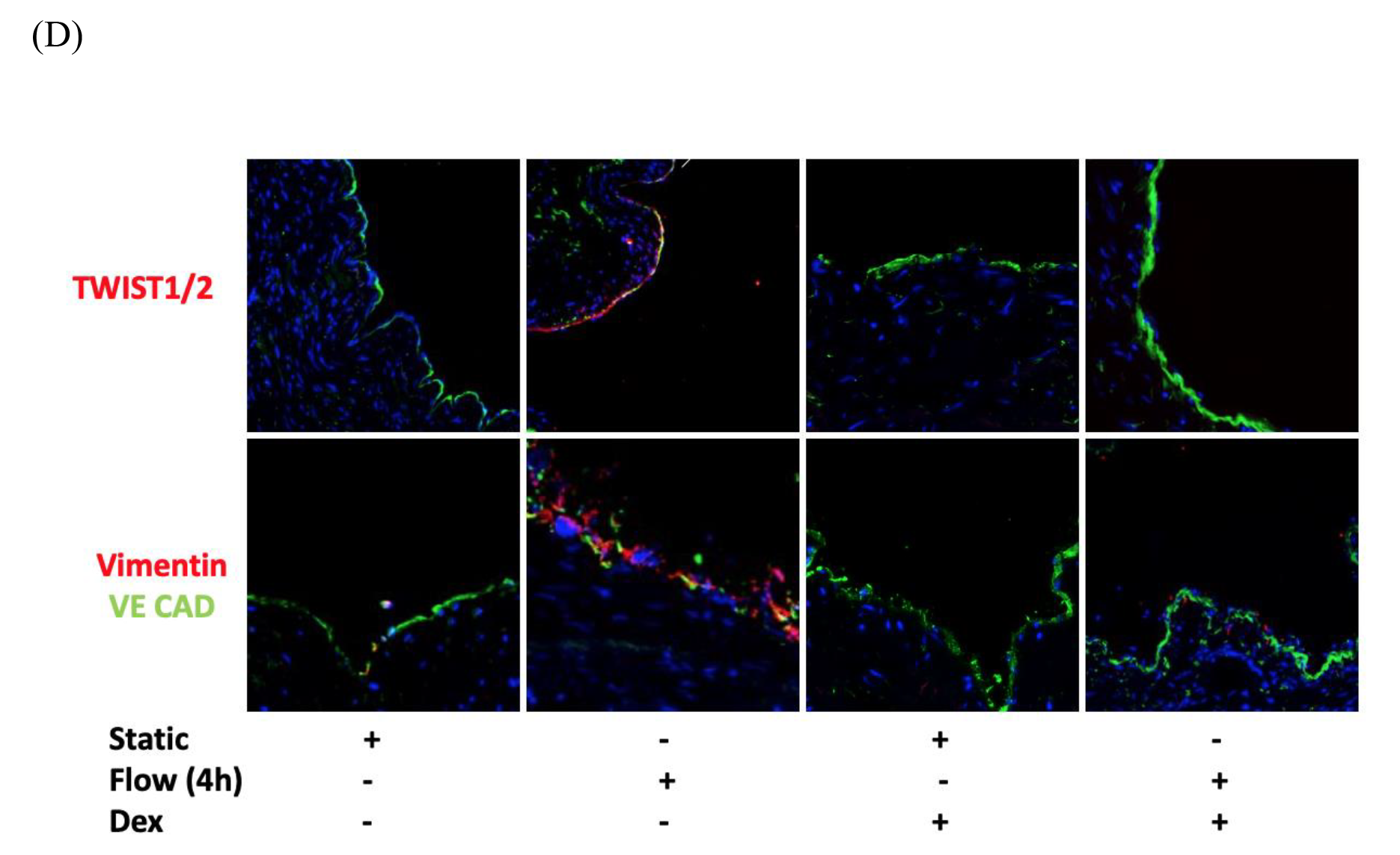
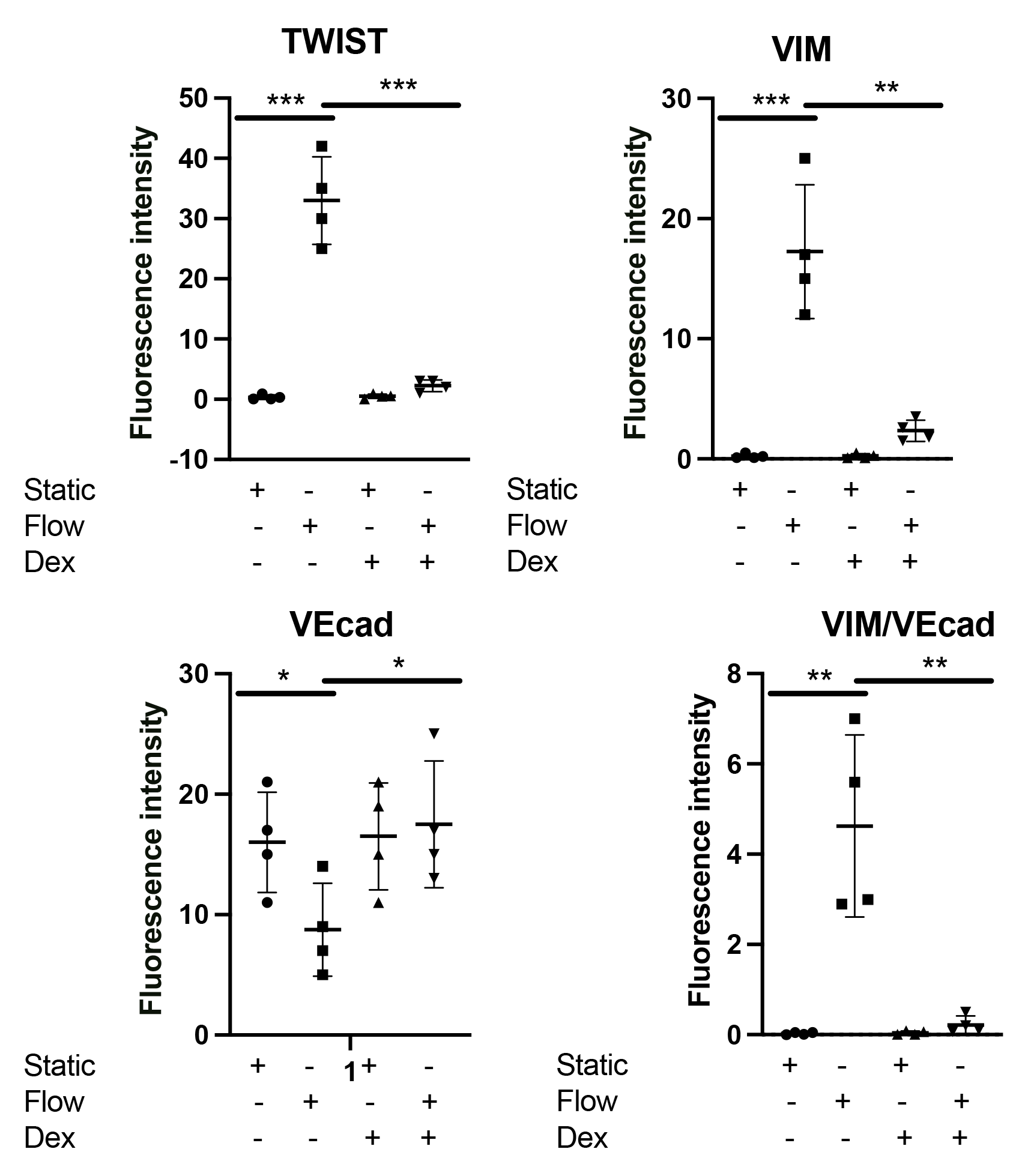
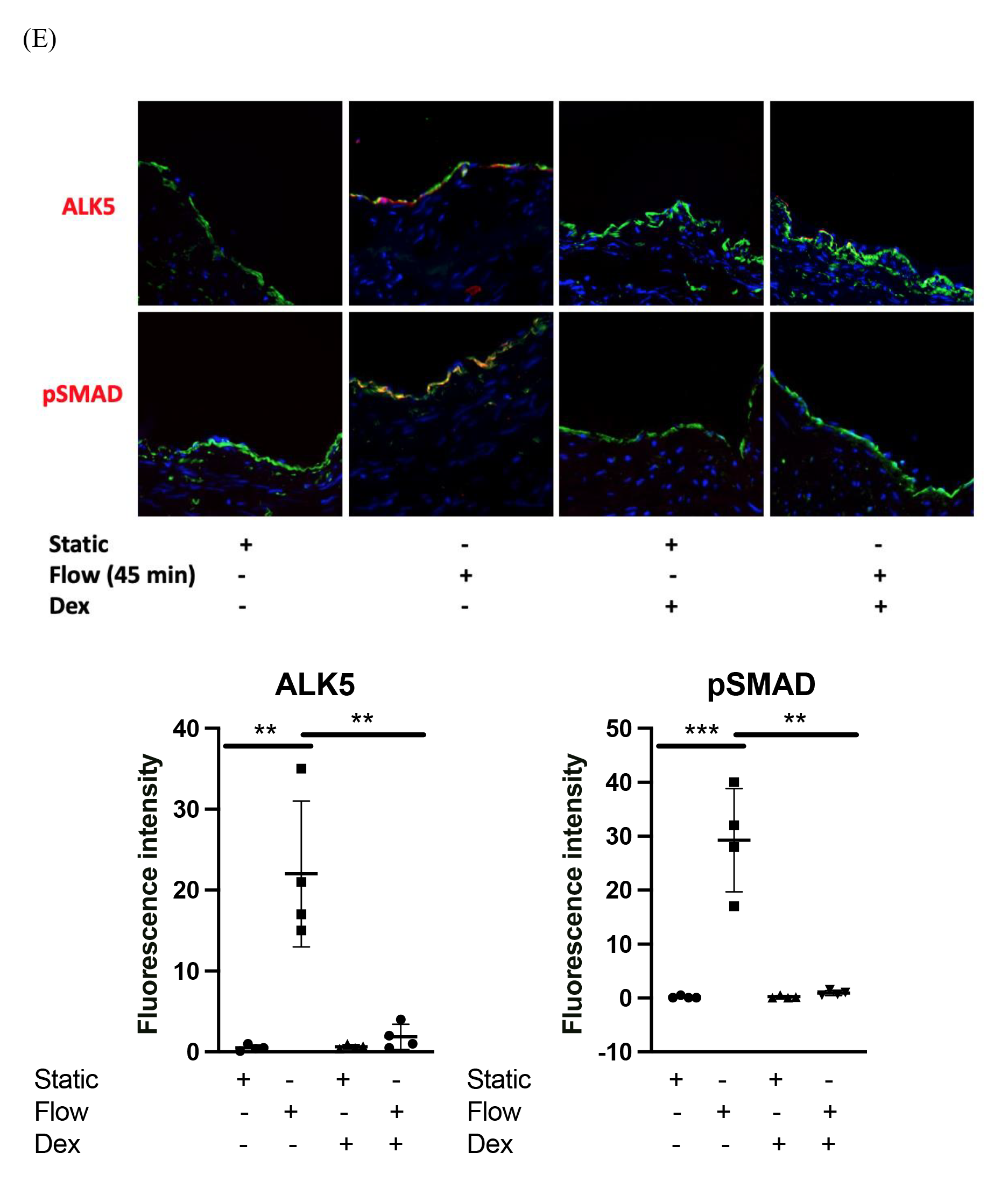
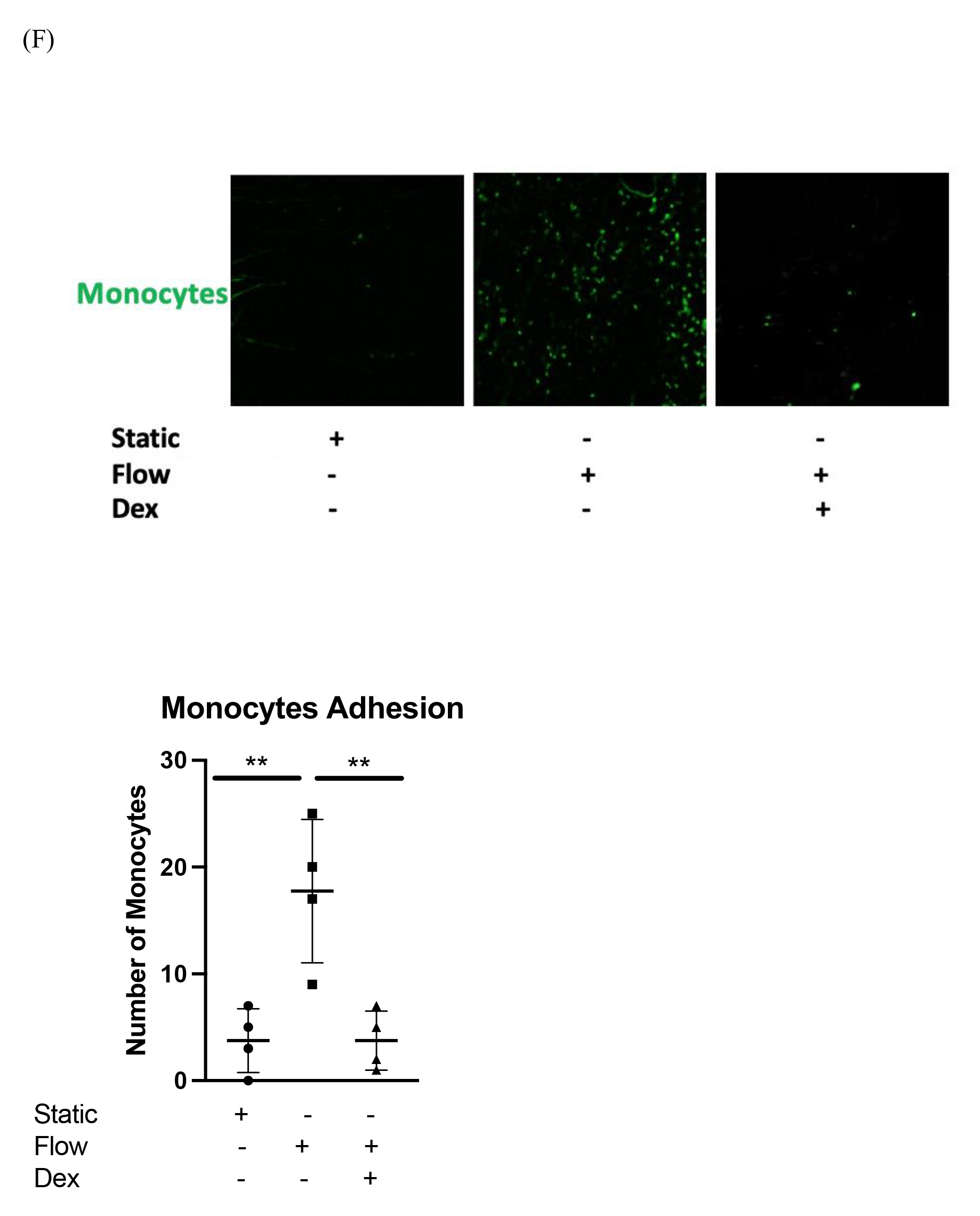
Pretreatment of human long saphenous veins with dexamethasone suppressed EndMT activation by shear stress in EC. (A) LSV were either were pretreated with dexamethasone (10 μmol/L for 30 minutes) or remained untreated. They were then mounted on a perfusion apparatus and exposed to LSS for 4 hours or were maintained under static conditions as a control. Transcript levels of TWIST2 and TWIST1 from whole tissue were measured by comparative RT-PCR. Values from four independent experiments and mean values are shown. (B) Transcript expression and levels of TWIST2 and TWIST 2 at 4 hours were assessed using RNASCope and probes specific for TWIST1 or TIWST2 genes (arrows indicating the expression of genes) quantified in multiple ECs, and averaged for each experimental group. Representative images, values from four independent experiments, and mean values are shown. (C) LSV were either were pretreated with dexamethasone (10 μmol/L for 30 minutes) or remained untreated. They were then mounted on a perfusion apparatus and exposed to LSS for 4 hours or were maintained under static conditions as a control. Transcript levels of VEcad and VIM from whole tissue were measured by comparative RT-PCR. Values from four independent experiments and mean values are shown. (D) TWIST1/2, VIM and VEcad expression levels were assessed at 4 hours by immunofluorescence staining with specific antibodies, quantified in multiple ECs, and averaged for each experimental group. Representative images, values from four independent experiments, and mean values are shown. (E) ALK5 and Psmad 2/3 expression levels were assessed at 45 minutes by immunofluorescence staining with specific antibodies, quantified in multiple ECs, and averaged for each experimental group. Representative images, values from four independent experiments, and mean values are shown. (F) LSV were either were pretreated with dexamethasone (10 μmol/L for 30 minutes) or remained untreated. They were then mounted on a perfusion apparatus and exposed to LSS for 4 hours or were maintained under static conditions as a control. Following exposure to flow, LSV sections were immediately prepared *en face* and incubated with 1×10^6^ Calcein-labelled monocytes per mL for 15 min then washed and imaged immediately and the total number of monocytes was calculated using CellProfiler software.

Interestingly, when we compared the role of low flow on the activation of LSV ECs and noted that low flow did not trigger any EndMT changes in LSV *ex-vivo* (Supp Figure. 3E). Thus, we conclude that a brief pretreatment with Dex, can suppress EndMT activation in LSV ECs in response to shear stress by suppressing the TGFβ /SMAD1 pathway.

## Discussion

Veins grafted into the arterial circulation are exposed to a sudden increase in blood flow. It is recognised that haemodynamics can influence the inflammatory process by controlling leukocyte margination and adhesive interactions and by generating shear stress, which alters EC physiology. (4-6, 37) We have previously shown that that venous and arterial ECs respond differently to shear stress suggesting that the susceptibility of veins and arteries to the inflammatory process is regulated by an interaction between local haemodynamics and stable vessel-specific EC phenotypes. (7) We also demonstrated that high but not low shear stress is associated with such proinflammatory responses in venous ECs. (8) In this study, we looked at the impact of exposing venous EC/ veins to arterial haemodynamics on the activation of EndMT and the pathway involved. EndMT can be stimulated by many factors with the most common of these are the TGFβ and BMP family of growth factors. (38-40) TGFβ1 binds to a complex of receptors that include type II receptor (TβRII) and type I receptor activin-like kinase 5 (ALK5), which promotes signalling through SMAD2/3 (41) whereas BMP2 and BMP4 primarily signal through the ALK2 receptor. (42, 43)

Since the presence of TGFβ in the neointima of animal models was reported (44), experimental studies have shown that upregulation of TGFβ or addition of exogenous TGFβ resulted in increased intimal thickening or neointima formation. (45-47) Moreover, many groups demonstrated that that TGFβ1 is upregulated early after injury. (48-50) Such upregulation can separately activate both SMAD1/5- and SMAD2-mediated pathways which can play an essential role in the development of IH in vein grafts. (51)

Cooley et al. using in vivo murine cell lineage-tracing models, we show that endothelial-derived cells contribute to neointimal formation through EndMT, which is dependent upon early activation of the SMAD2/3-Slug signalling pathway. (52) Histological examination of post-mortem human vein graft tissue corroborated the changes observed in our mouse vein graft model, suggesting that EndMT is functioning during human vein graft remodelling.

Our work supports and expand on what has been previously reported. We first examined the effects of flow on the expression of the EndMT markers *in-vitro*. We observed that acute exposure to high shear stress was associated with rapid activation of ALK5, pSMAD 2/3, activation of TWIST 2 and TWIST 1 transcription factors and changes in EC supporting phenotype changes. We confirmed that these EndMT are mediated by ALK5 by using a specific inhibitor that was able to suppress the impact of acute shear on EC. Next, we showed that the silencing of TWIST 1 and TWIST 2 transcription factors can suppress EndMT changes in EC confirming their role. Based on our previous work, we showed that a brief pre-treatment of EC with Dex can suppress the activation OF EndMT *in-vitro*. Next, we carried out spatial cell sequencing of LSV sections exposed to arterial flow and showed that activation of different proinflammatory mediators and TWIST 1 & 2 in response to acute arterial flow based on clustering which identified a subset of cells that has both EC and SMC characteristics. In this cluster, we noted that TWIST 2 was significantly activated by flow suggesting that these cells are at intermediate stage of EndMT or what has been described previously as the hybrid cells expressing both EC and SMC specific molecular markers and most likely representing an intermediary phenotype between endothelial cells and mesenchymal/myofibroblastic cells. (53) We validated these untargeted results *ex-vivo* using RT-QPCR, RNAscope and immunostaining. This showed that the exposure of LSV to acute arterial flow *ex-vivo* can trigger EndMT in EC. Moreover, we demonstrated that a brief pretreatment of LSV with Dex can suppress these changes. The oral administration of a non-specific inhibitor of TGFβ biosynthesis in patients after PCI failed to show significant improvement, and was associated with significant adverse effects (54) considering that TGFβ signalling pathway is very complex and mediates several functions in other organs, thus the ability to use local/ topical therapeutics that can target pathways in specific tissue without exposing patients to the adverse effects of systemic usage is more attractive alternative. Glucocorticoids are known to inhibit TGFβ production and modulate the activities of different members of the SMAD family and EndMT. (15, 16, 18) Here, we provide proof of principle that brief pretreatment with Dex can prevent veins from exhibiting an inflammatory phenotype in response to arterial shear stress. Our findings suggest that the known beneficial effects of Dex treatment in the prevention of intimal hyperplasia may involve suppression of TGFβ /SMAD EndMT in addition to suppressing vascular inflammation via MAPK pathway. They also suggest that a brief pretreatment with Dex may be sufficient to protect veins from early inflammatory responses to grafting. Perioperative treatment of veins may therefore provide an attractive novel therapeutic strategy to reduce graft inflammation and allowing veins to adapt to their new environment while avoiding the undesirable side effects associated with systemic glucocorticoid treatment.

Although we focused on ALK5/ SMAD pathway in this study, evidence suggests that Glucocorticoids are effective in inhibiting bone morphogenetic proteins expression such as BMP2 (55) thus it will be interesting in future studies to specifically address EndMT in LSV in relation to BMPs specifically BMP2 based on our sequencing data which showed significant BMP2 but no bmp4 activation in the whole tissue sequencing but not in the EC/SMC subset based on our network analysis which may suggest the it can play a role at different stage of phenotype changes. Additionally addressing the role of NF-kB in triggering EndMT via the activation of transcription factors such as TWIST 1&2 will be very interesting taking into account spatial data network analysis and the links between NF-kB, inflammation and TWIST 2.

### Source of Funding

This study was funded by the Van Geest foundation charitable fund and the BHF accelerator award.

### Disclosures

None.

## Data Availability

All data produced in the present study are available upon reasonable request to the authors

---

**Supp Figure. 1.**
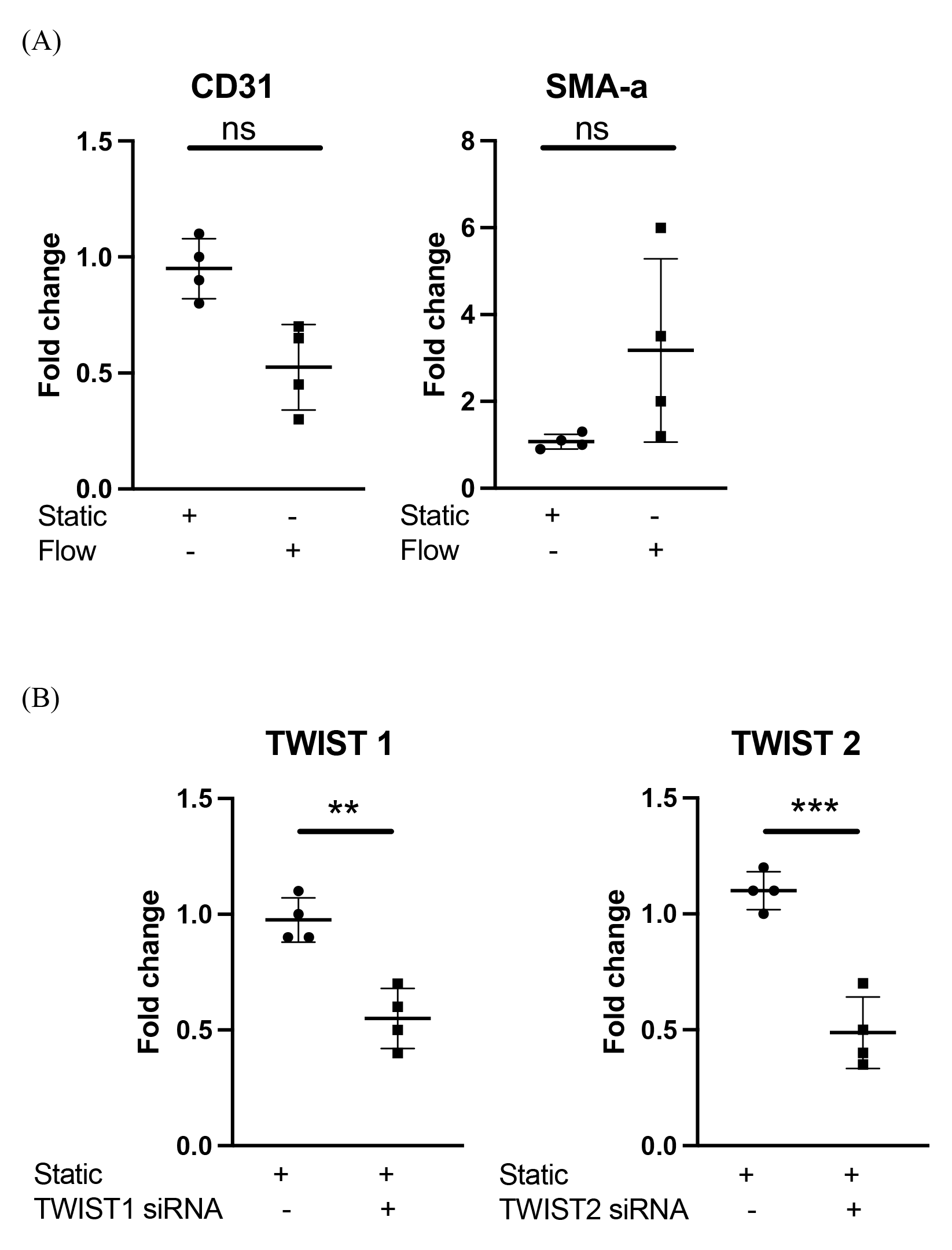
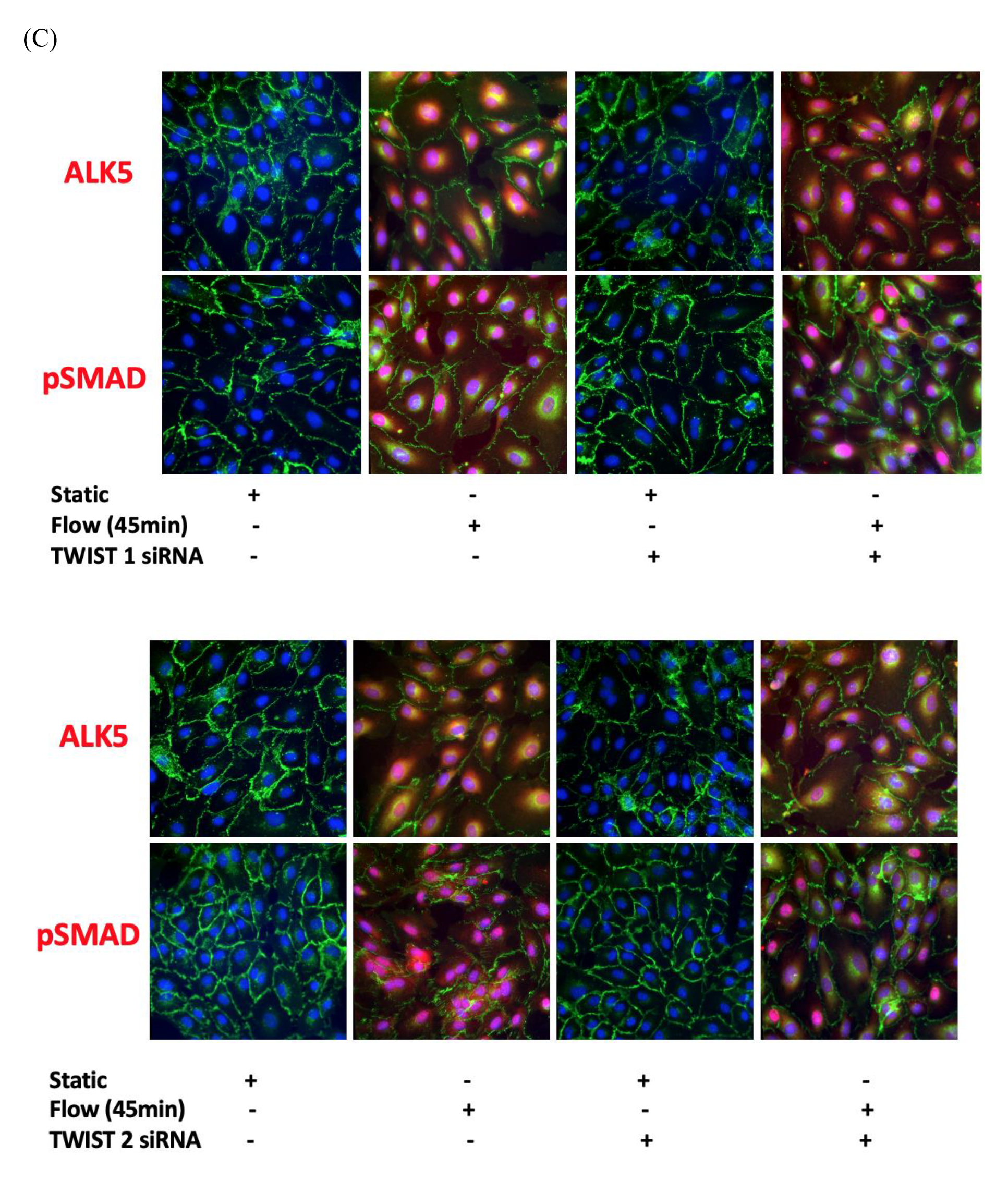
(A) HUVECs were cultured under static conditions or exposed to LSS for 4 hours. Transcript levels of CD31 and SMA were measured by comparative RT-PCR. Values from four independent experiments and mean values are shown. (B) HUVECs were cultured under static conditions or exposed to LSS for 4 hours. Transcript levels of TWIST1 and TWIST2 were measured by comparative RT-PCR. Values from four independent experiments and mean values are shown. (C) HUVECs were transfected with TIWST1 or TWIST2–specific siRNA or with a scrambled control (Scr). expression levels were assessed for ALK5 and Psmad 2/3 assessed at 45 minutes by immunofluorescence staining with specific antibodies, quantified in multiple ECs, and averaged for each experimental group. Representative images, values from four independent experiments.

**Supp Figure. 2.**
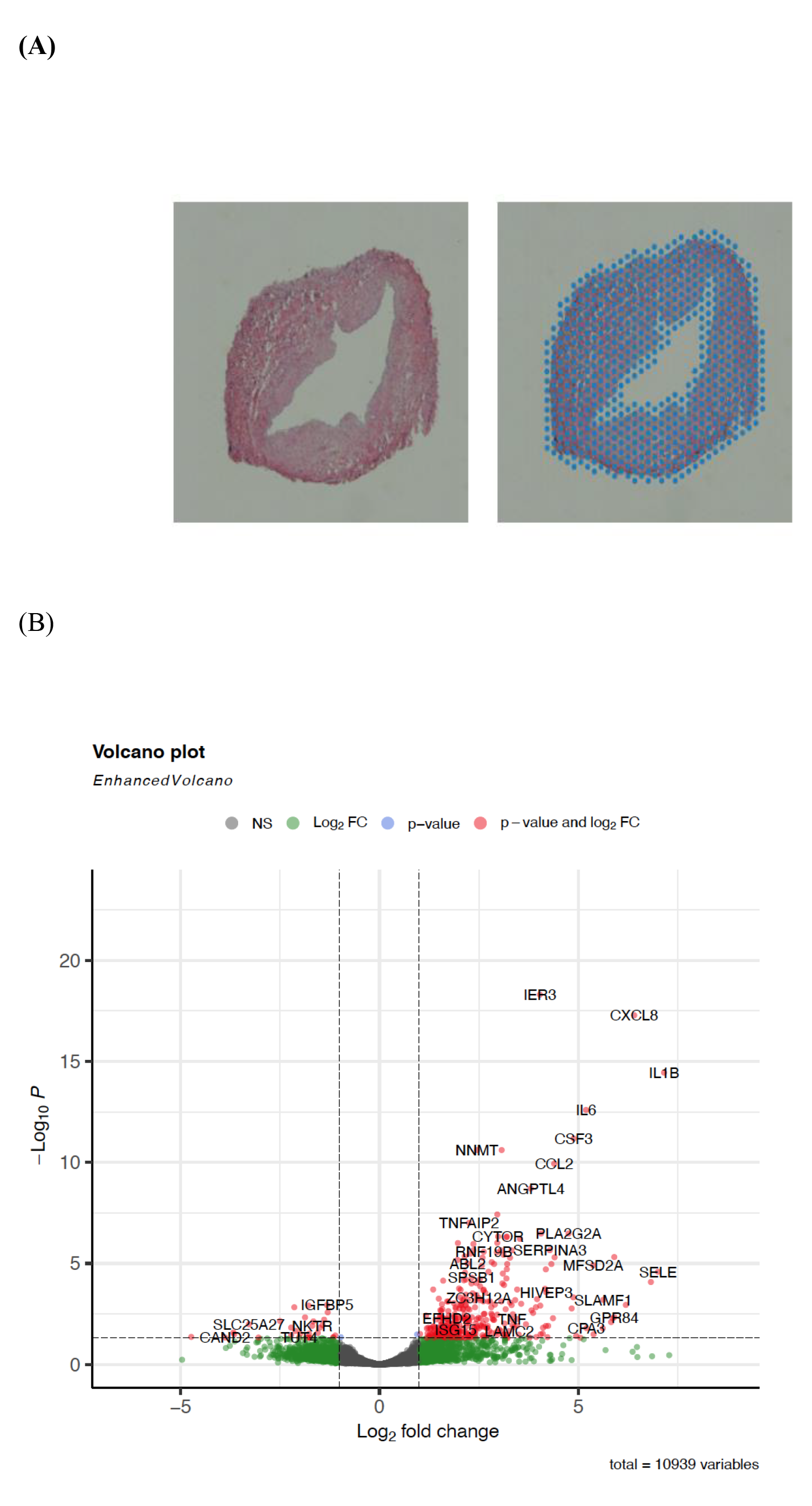
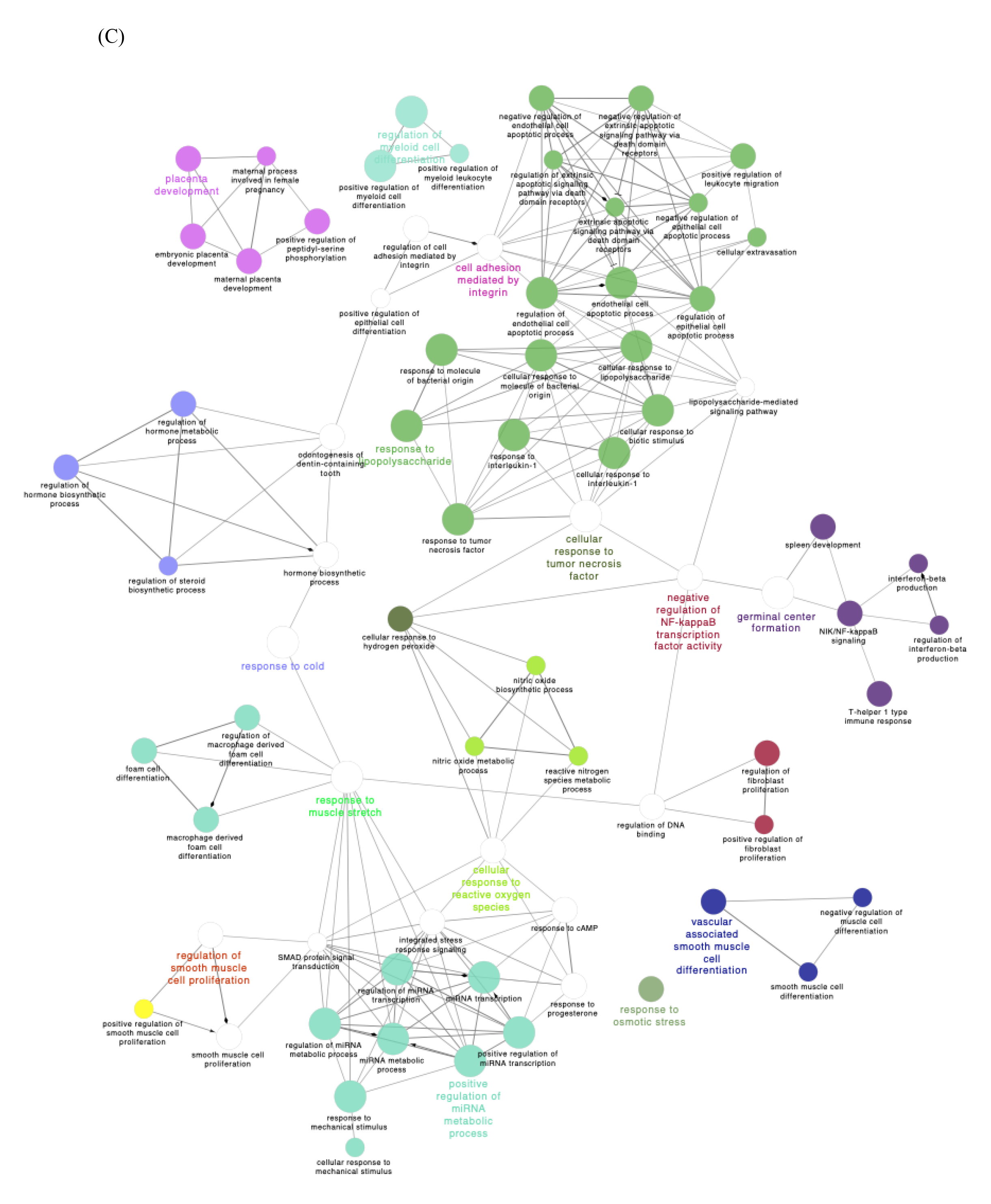
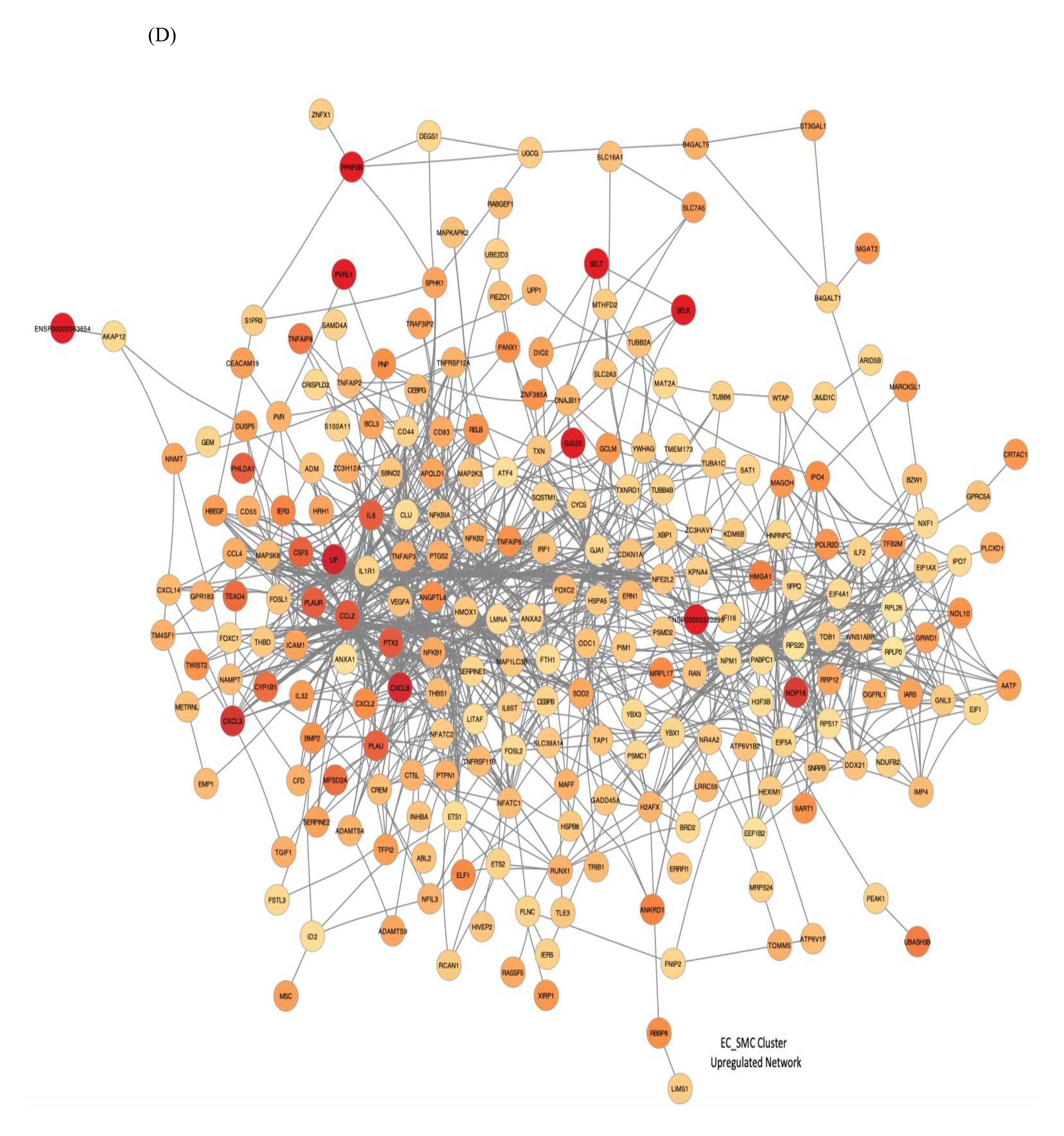
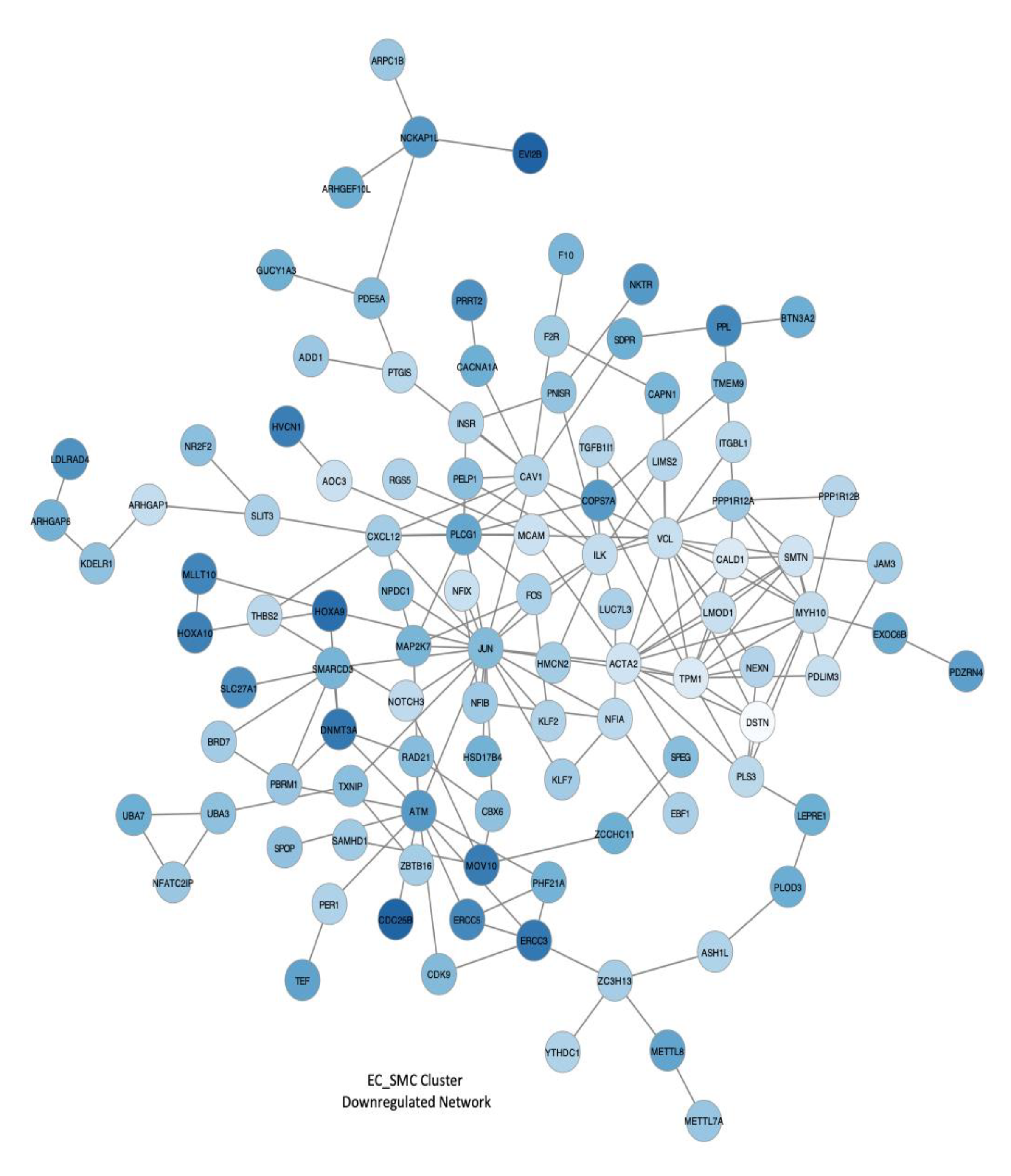
(A) representative images of LSV tissue section (10um) microscopy image and its associated 10X Visium spot coverage grid overlay, indicating the manually selected RNA capture area utilised for analysis. (B) Volcano plot depicting all differentially expressed genes (DEGs) between the control samples and samples exposed to 4 hours arterial haemodynamics (n = 4 each). Genes identified in red exceed the p-value and log2FC thresholds for significance. NS = not significant. (C) Global pathway analysis of the whole tissue of all significant differentially expressed genes (adjusted p-value < 0.05) with the top 30 connections presented. Enrichment is scaled based on significance (-log10 p_value) and visually ranked based on intersection size (overlap of DEGs with enriched pathways). (D) EC/SMC cluster network analysis of all significant differentially upregulated and downregulated genes (adjusted p-value < 0.05) in samples exposed to *ex vivo* arterial haemodynamic conditions. Samples are coloured on a red-grey gradient based on logFC values. Direct connections illustrate known direct interactions.

**Supp Figure. 3.**
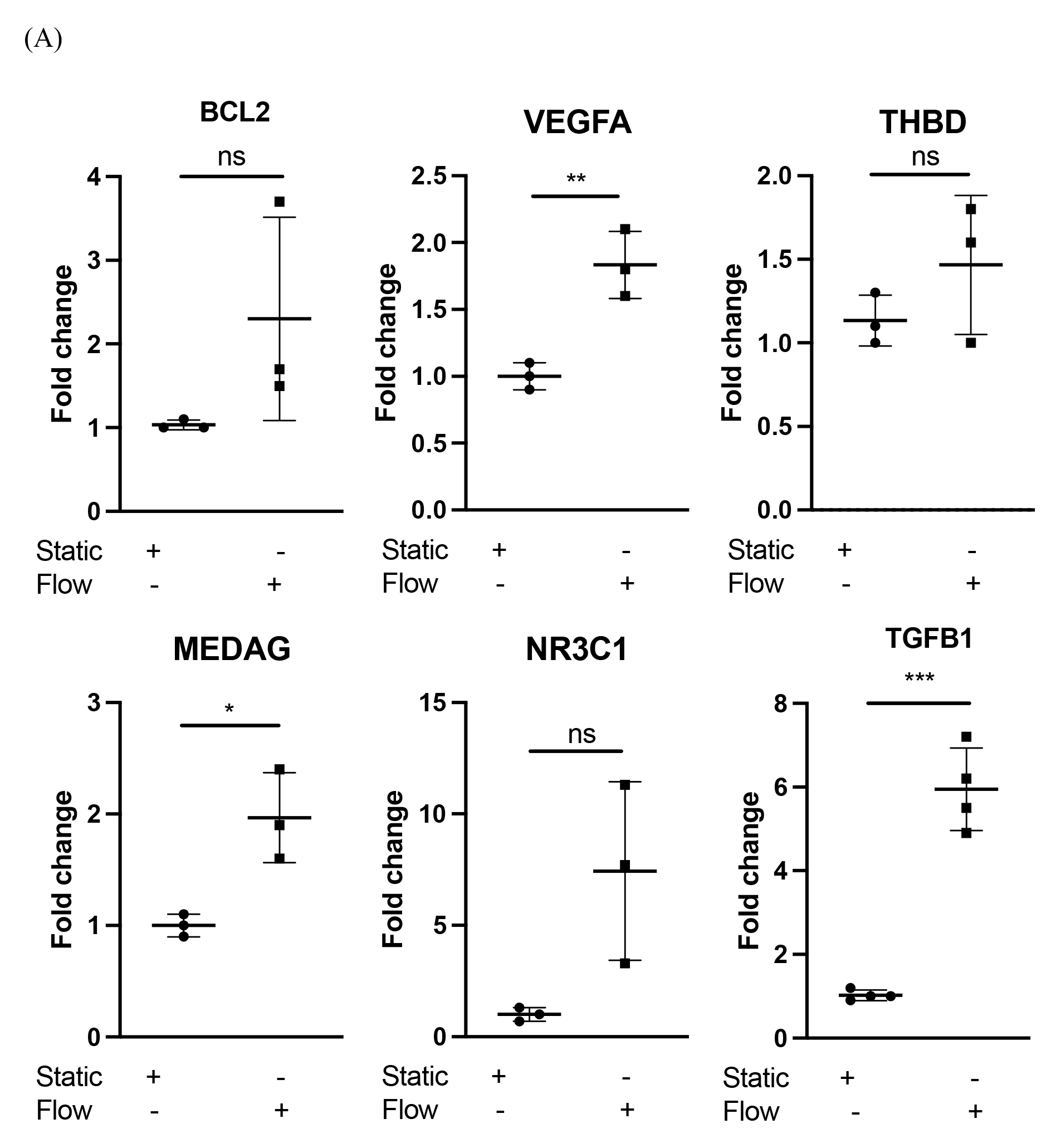
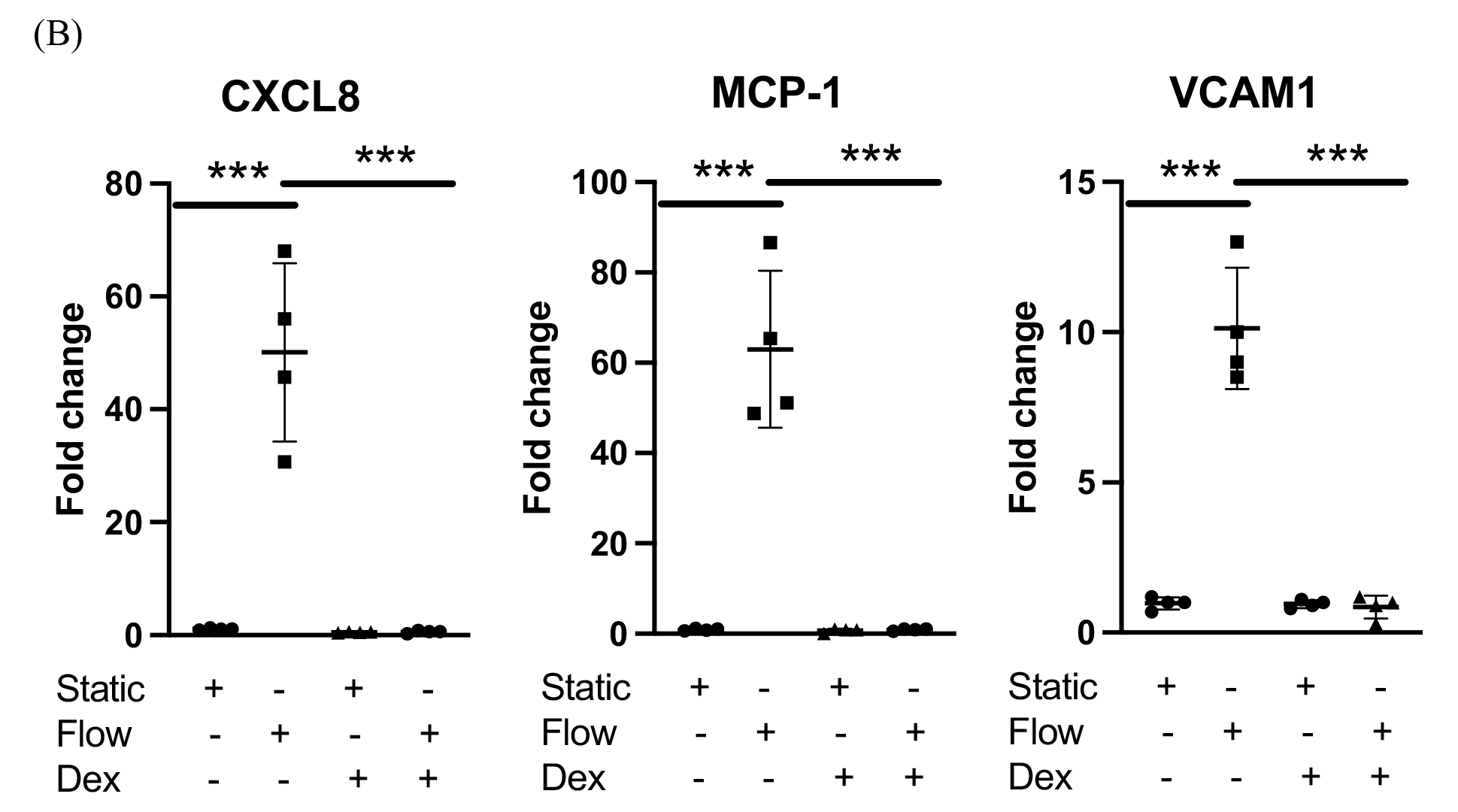
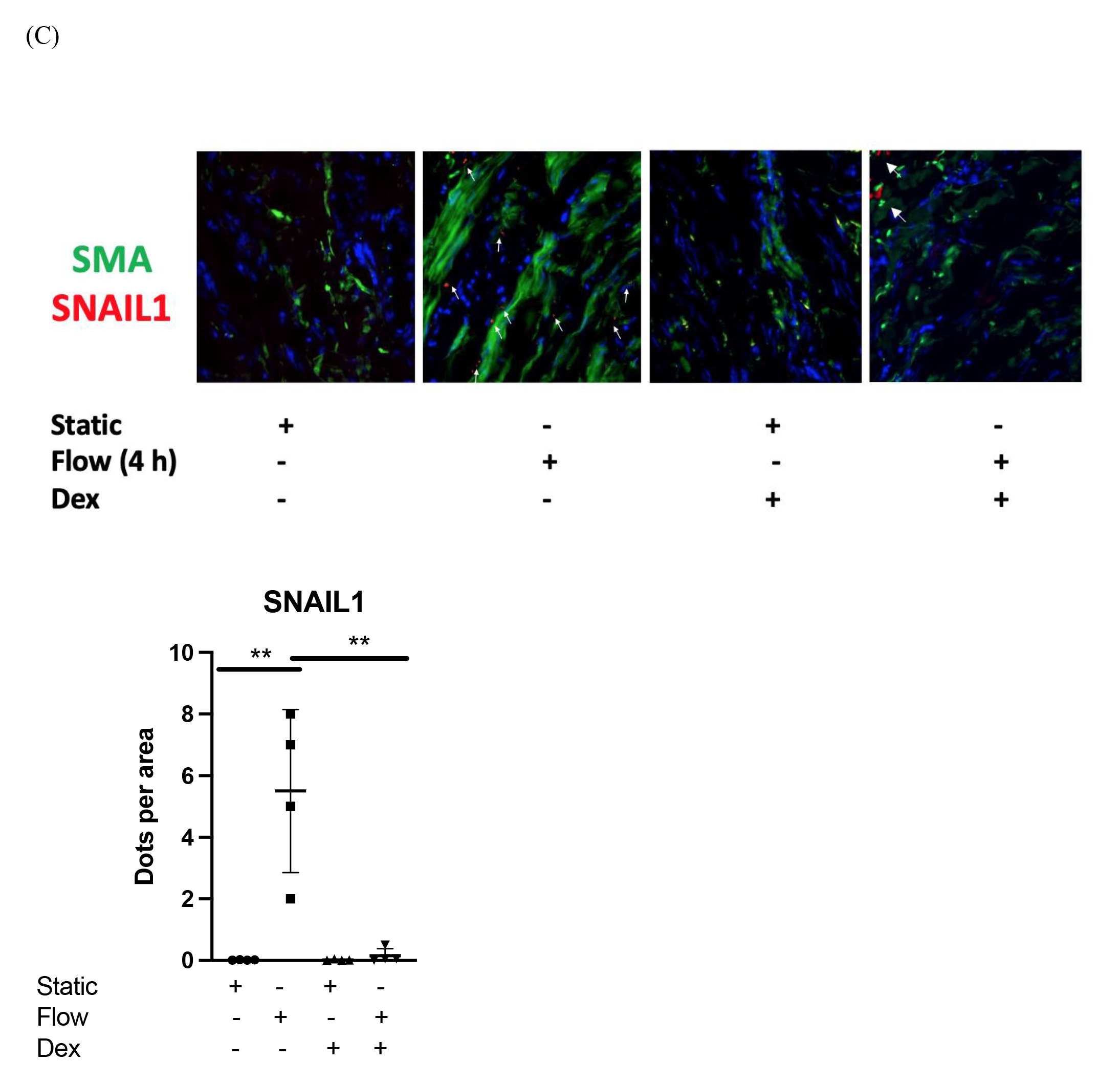
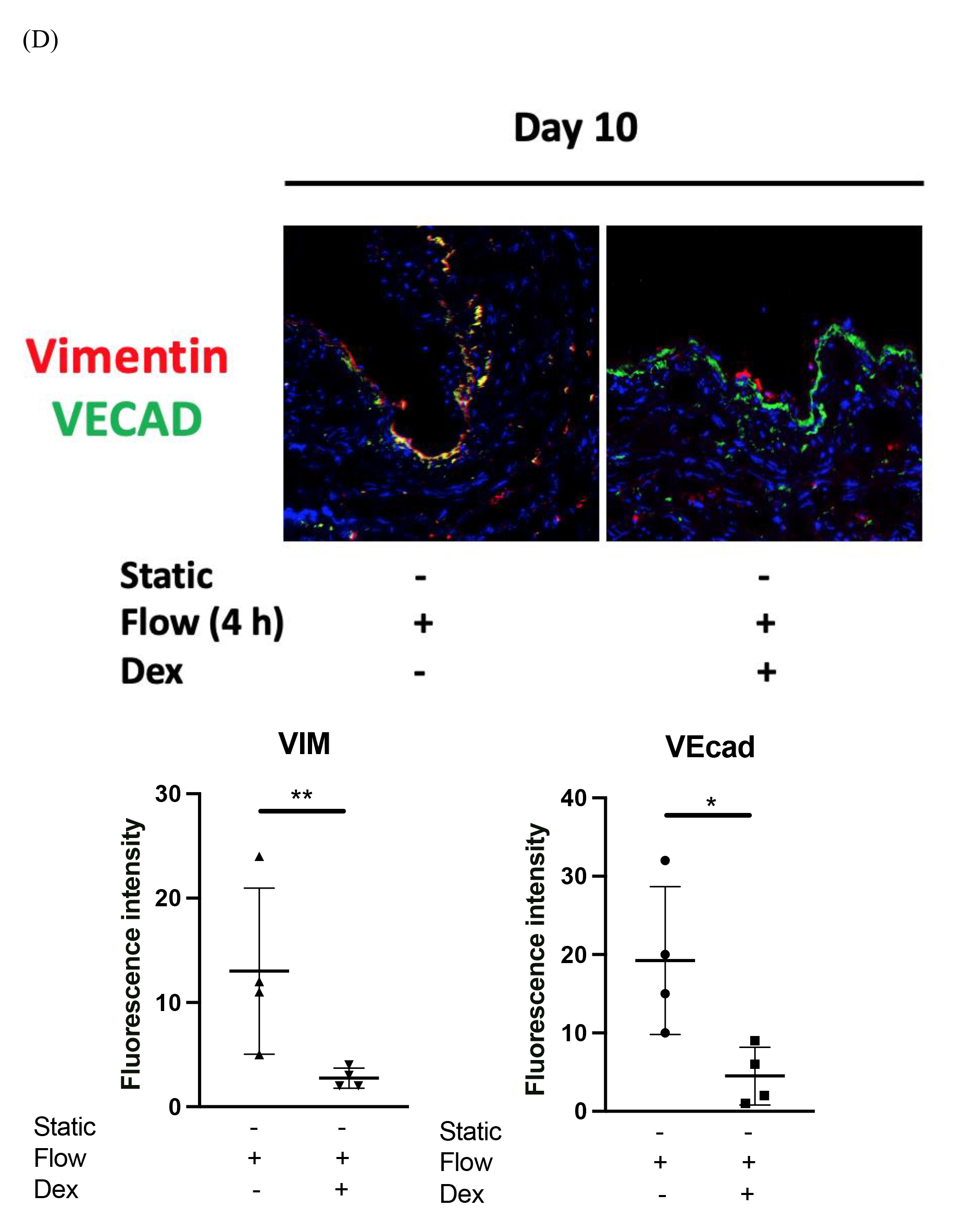
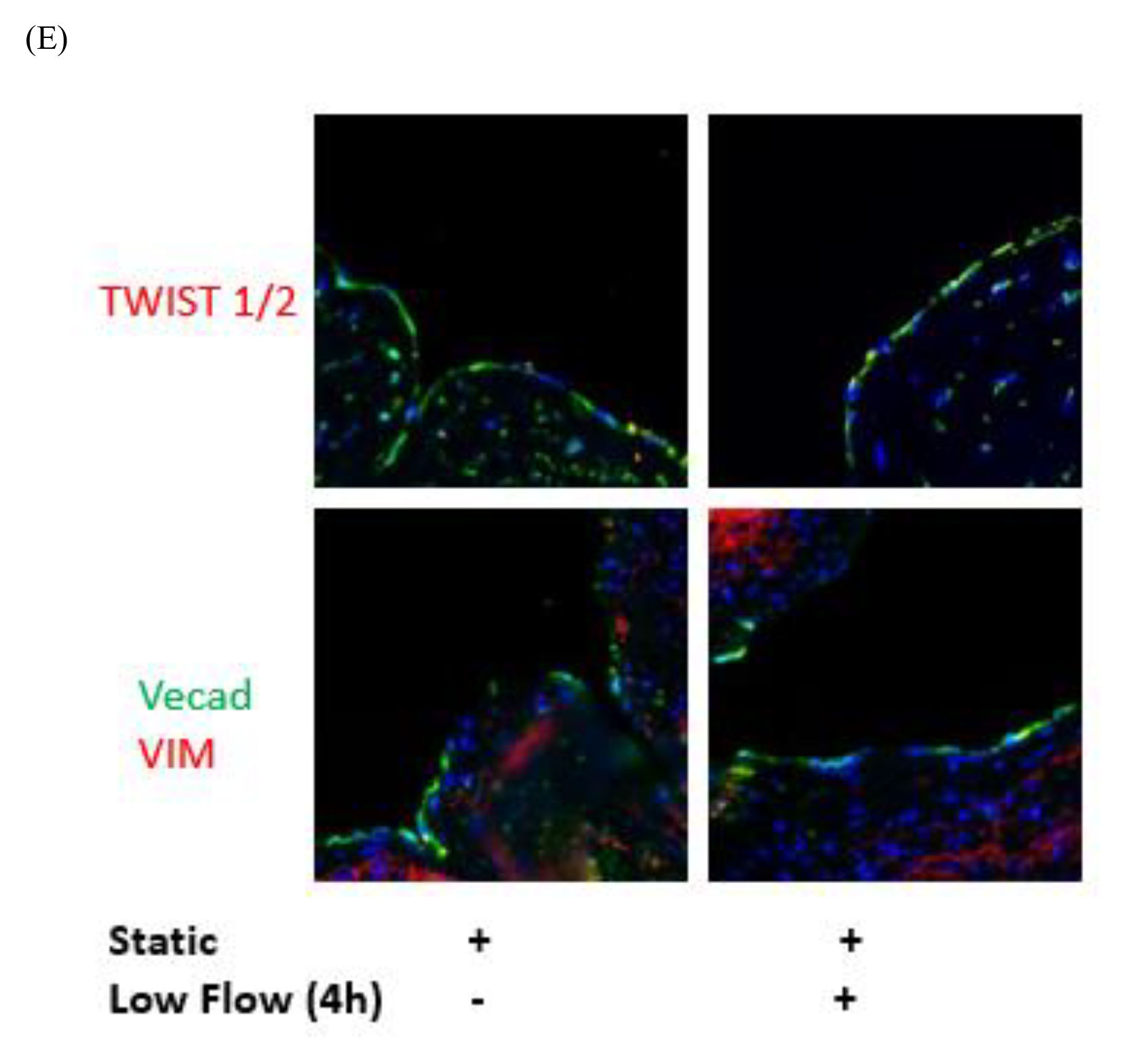
(A) LSV were either were pretreated with dexamethasone (10 μmol/L for 30 minutes) or remained untreated. They were then mounted on a perfusion apparatus and exposed to LSS for 4 hours or were maintained under static conditions as a control. Transcript levels of CXCL8, MCP-1 and VCAM1 from whole tissue were measured by comparative RT-PCR. Values from four independent experiments and mean values are shown. (B) LSV were either were pretreated with dexamethasone (10 μmol/L for 30 minutes) or remained untreated. They were then mounted on a perfusion apparatus and exposed to LSS for 4 hours or were maintained under static conditions as a control. Transcript levels of genes of interest from the spatial sequencing data were validated by comparative RT-PCR. Values from four independent experiments and mean values are shown. (C) Transcript expression and levels of SNAIL1 at 4 hours were assessed using RNASCope and probes specific for SNAIL1 gene (arrows indicating the expression of genes) quantified in multiple ECs, and averaged for each experimental group. Representative images, values from four independent experiments, and mean values are shown. (D) LSV were either were pretreated with dexamethasone (10 μmol/L for 30 minutes) or remained untreated. They were then mounted on a perfusion apparatus and exposed to LSS for 4 hours or were maintained under static conditions as a control. Vein segments were next cultured in a culture well for 10 days and expression levels for VIM and VEcad was assessed by immunofluorescence staining with specific antibodies, quantified in multiple ECs, and averaged for each experimental group. Representative images, values from four independent experiments. (E) LSV were either were mounted on a perfusion apparatus and exposed to low shear for 4 hours or were maintained under static conditions as a control TWIST1/2, VIM and VEcad expression levels were assessed at 4 hours by immunofluorescence staining with specific antibodies, quantified in multiple ECs, and averaged for each experimental group. Representative images from four independent experiments are shown.

